# Longitudinal 7T MRS Study of Glutamate and GABA Dynamics in Alzheimer’s Disease Progression: From hyper- to hypoexcitation

**DOI:** 10.64898/2026.02.04.26345353

**Authors:** Laura Göschel, Andrea Dell’Orco, Semiha Aydin, Charlotte E. Teunissen, Patty L. Hoede, Jeanette Melin, Leslie Pendrill, Sebastian Roemer-Cassiano, Nicolai Franzmeier, Agnes Flöel, Péter Körtvélyessy, Ariane Fillmer

## Abstract

Functional neuroimaging studies suggested a biphasic trajectory of neuronal activity in Alzheimer’s disease (AD), with early hyperactivity followed by later hypoactivity. However, the underlying neurochemical mechanisms in humans remain unclear. Animal studies suggested that amyloid-beta (Aβ) causes intrasynaptic glutamate increases through impaired astrocytic clearance. This study aimed to build a mechanistic bridge between findings from human neuroimaging studies and preclinical models by providing in vivo measurements of glutamate, GABA, and glutamine across the AD continuum using 7T magnetic resonance spectroscopy (MRS).

We acquired longitudinal data from cognitively normal (CN) individuals with and without Aβ pathology (CN Aβ-n=43, CN Aβ+ n=17), individuals with mild cognitive impairment (MCI Aβ+, n=20) or AD dementia (AD Aβ+, n=24). Over the course of up to 5 years, glutamate levels increased in the CN Aβ+ and MCI Aβ+ groups but decreased in the AD Aβ+ group. Consistent with a transient homeostatic response to glutamatergic hyperexcitation, GABA levels showed modest increases in both CN Aβ+ and MCI Aβ+ groups. Elevated plasma GFAP was associated with reduced glutamine, suggesting astrocytic dysfunction and impaired glutamate–glutamine cycling as contributors to early hyperexcitability. Together, our results provide the first direct demonstration in patients with AD that glutamate alterations underlie the biphasic trajectory of neuronal activity and that astrocytic glutamate regulation might be a potential therapeutic target.

## 1. Introduction

Recent neuroimaging studies indicated that changes in neuronal activity in individuals with Alzheimer’s disease (AD) pathology follow a biphasic trajectory, often described as an inverted U-shaped curve, characterized by an initial phase of hyperactivity (Gallego-Rudolf et al., 2024; Giorgio et al., 2024; Roemer-Cassiano et al., 2025; van Nifterick et al., 2023), followed by a later decline toward hypoactivity. This shift in neuronal activity was hypothesized to be driven primarily by amyloid-beta (Aβ) deposition (causing hyperactivity) and subsequent tau pathology and neuronal loss (causing hypoactivity, Gallego-Rudolf et al. (2024), for review Hector & Brouillette (2021), Targa Dias Anastacio et al. (2022)). Importantly, all of this is thought to occur before any cognitive deficits appear (Gallego-Rudolf et al., 2024). The mechanisms underlying the Aβ-induced hyperactivation, however, remain unclear.

Glutamate is the brain’s primary excitatory neurotransmitter (Sheldon & Robinson, 2007), while Gamma-aminobutyric acid (GABA) serves as the primary inhibitory neurotransmitter (Govindpani et al., 2017). Although AD is not classically defined as a neurotransmitter disorder, disruptions in glutamatergic and GABAergic signaling are increasingly recognized as key components of AD pathophysiology: Several studies of mouse models have demonstrated that Aβ pathology, especially Aβ oligomers, stimulate glutamate release, impair post-synaptic glutamate uptake, and disrupt astrocytic clearance leading to increased glutamate levels in the synaptic cleft (Rudy et al., 2015; Zott & Konnerth, 2023). Specifically, an Aβ-related impairment of the glutamate-glutamine cycle in astrocytes might play an essential role in increases of glutamate levels (Zott & Konnerth, 2023). Glutamatergic homeostasis critically depends on astrocytic uptake from the synaptic cleft and recycling to glutamine via the glutamate–glutamine cycle (Rudy et al., 2015). The resulting prolonged glutamate receptor activation is believed to 1) increase the risk of seizures, observed in up to 22% of AD patients (Barbour et al., 2024; Vossel et al., 2013), 2) enhance exocytotic tau release and propagation (Mazzo et al., 2022; Roemer-Cassiano et al., 2025), and 3) contribute to synaptic loss, neuronal death, and consequently, memory deficits (Rudy et al., 2015). Compared with glutamate, the interaction between the GABAergic system and AD pathology remains poorly understood, with studies reporting divergent findings (Carello-Collar et al., 2023; Govindpani et al., 2017). To date, the suggested interplay between Aβ pathology and neurotransmitter systems has not been demonstrated in vivo in humans, although pharmacologic interventions to modulate glutamatergic and GABAergic excitation may be promising therapeutic targets for AD (Scaduto et al., 2023; Sheldon & Robinson, 2007).

Magnetic Resonance Spectroscopy (MRS) is a non-invasive technique for in vivo measurements of the concentration of specific biochemicals. Although widely used to study brain metabolites, MRS has rarely been applied to investigate alterations in glutamate and GABA in AD. One major challenge is that the glutamate resonance overlaps with that of glutamine, which is the precursor of both glutamate and GABA (McKiernan et al., 2023). Among studies that quantified absolute glutamate, most reported lower glutamate levels in AD compared with healthy controls (Fayed et al., 2011; Rupsingh et al., 2011; Wong et al., 2020), whereas the one study conducted at ultra-high-field strength 7T found no group differences (Marjańska et al., 2019). In individuals with mild cognitive impairment (MCI), often considered a prodromal stage of AD, findings have been similarly inconsistent, with reports of reduced glutamate (Wong et al., 2020; Zeydan et al., 2017; Zeydan & Kantarci, 2021) as well as unchanged levels (Fayed et al., 2011; Rupsingh et al., 2011). Notably, the only two 7T MRS studies that included MCI participants also reported lower glutamate concentrations or glutamate/creatine ratios (Oeltzschner et al., 2019; Wong et al., 2020). While reduced glutamate in advanced AD may reflect neuronal and synaptic loss, similar reductions observed in MCI are challenging to reconcile with preclinical evidence of Aβ-driven glutamatergic hyperexcitability in early disease stages. This discrepancy may, at least in part, arise from methodological limitations of prior studies, and a confirmation with longitudinal, high-resolution MRS studies with more refined disease staging is needed.

Unlike glutamate, GABA is found at low concentrations in the brain, and its quantification by MRS is technically challenging due to spectral overlap with more abundant metabolites, including N-acetylaspartate (NAA), Creatine (Cr), glutamate, and glutamine (McKiernan et al., 2023). Additionally, macromolecule contamination can account for up to 60% of the GABA signal at conventional field strengths, resulting in the composite measure GABA+ (Deelchand et al., 2019). Despite these limitations, clinical MRS studies have consistently reported lower GABA+ in MCI patients when compared to healthy controls (Fu et al., 2023; Hone-Blanchet et al., 2022; Riese et al., 2015), with one exception reporting no differences in either MCI or AD (Huang et al., 2017). Ultra-high-field MRS offers substantial advantages for GABA quantification, including improved signal-to-noise ratio, shorter acquisition times that reduce motion artefacts, and reduced macromolecule contamination (Deelchand et al., 2019; Dell’Orco et al., 2024). To date, the only two 7T MRS studies reported reduced GABA concentrations in both MCI (Oeltzschner et al., 2019) and AD (Marjańska et al., 2019), consistent with preclinical evidence (Carello-Collar et al., 2023). However, it remains unclear whether GABA changes progress in parallel with disease severity, and longitudinal investigations are needed to characterize the temporal dynamics of inhibitory neurotransmission in AD.

Also glutamine is difficult to quantify by MRS due to its relatively low concentration and substantial spectral overlap with glutamate (McKiernan et al., 2023). For this reason, these metabolites are usually quantified together as Glx (glutamate + glutamine; McKiernan et al., 2023). Nevertheless, absolute glutamine levels measured at 7T were reported to be statistically unchanged in AD (Marjańska et al., 2019). Whether glutamine levels correlate with astrocytic reactivity or dysfunction, particularly across disease stages, has not yet been systematically investigated.

Previous human MRS studies have not been able to confirm a role for glutamate levels in the hyperactivity observed in early AD pathology, likely because ultra-high-field (7T) MRS studies with improved spectral resolution remain scarce, and most investigations have relied on cross-sectional group comparisons rather than individual longitudinal trajectories. To date, MRS studies of glutamate and GABA in AD have provided only static, usually group-level snapshots. Such designs cannot capture individual temporal changes, which are particularly relevant given the substantial heterogeneity in disease progression, resulting in overlap between cognitively normal, MCI, and AD groups. We argue that longitudinal high-field MRS studies are therefore needed to track glutamate and GABA over time, and to distinguish early excitotoxic phases from later stages dominated by neuronal loss. Understanding these stage-dependent dynamics could clarify disease mechanisms, identify optimal therapeutic windows, and inform the development of interventions that modulate glutamatergic and GABAergic transmission.

In this study, we aimed to build a mechanistic bridge between earlier neuroimaging studies of humans and glutamate-related studies of animal models, by applying longitudinal 7T MRS in cognitively normal individuals with and without Aβ pathology (CN Aβ-, n = 43, CN Aβ+, n = 17), MCI Aβ+ (n = 20), and AD Aβ+ (n = 24). We characterized glutamate and GABA trajectories in relation to disease progression and investigated a possible involvement of astrocytic reactivity in the glutamatergic upregulation. We specifically hypothesized that 1) glutamate levels are elevated in the early presence of Aβ pathology (CN Aβ+ and MCI Aβ+) and decrease in later stages of AD progression (AD Aβ+), 2) GABA decreases with disease progression and 3) astrocytic dysfunction (as indicated by elevated plasma glial fibrillary acidic protein (GFAP) levels) is associated with elevated glutamate levels through an impaired glutamate-glutamine metabolism.

## 2. Methods

### 2.1 Study Design and Participants

The present study used data from the projects 15HLT04 NeuroMET (2016-2019) and 18HLT09 NeuroMET2 (2019-2022), both of which were designed to advance the diagnosis and management of neurodegenerative diseases through high-quality and standardized measurement methods. Participants were aged 55 to 84 and underwent up to four follow-up visits in addition to the baseline assessment. Stratification into the groups cognitively normal (CN), MCI, and dementia due to suspected AD was performed by an experienced team of neurologists, neuropsychologists, and researchers, based on neuropsychological test results and clinical evaluation, including cerebrospinal fluid (CSF) results when available. Exclusion criteria comprised a history of drug or alcohol abuse, eating disorders, and severe or untreated medical, neurological, or psychiatric diseases that could interfere with cognition. Positivity of Aβ pathology was defined by plasma biomarkers as described below. For the present statistical analyses, we excluded individuals without MRS data at any visit (n = 8), and individuals with inconsistent plasma biomarker profiles: classified as Aβ positive (Aβ+, “see Section 2.2 for biomarker classification”) at visit 1 but Aβ negative (Aβ-) at follow-ups (n = 3), and individuals with an MCI or AD diagnosis who were classified as Aβ-throughout all visits (n = 12). The main reasons for drop-out before the study completion were lack of ability to consent due to advanced AD (n = 16), lack of motivation (n = 22), other severe diseases (n = 7), and death (n = 2).

This resulted in a final study sample of 104 individuals of the groups CN Aβ-(n = 43), CN Aβ+ (n = 17), MCI Aβ+ (n = 20), and AD Aβ+ (n = 24). Each study visit comprised a standardized medical interview, neurological examination, and extensive neuropsychological testing. At visit 1 and selected follow-up visits, participants additionally underwent blood collection, 7T structural and fMRI, and MRS. All participants were White and native German speakers, and they provided written informed consent before participating in the study. The study was approved by the ethics committee of Charité University Hospital (EA1/197/16 and EA2/121/19) and conducted in accordance with the Declaration of Helsinki. All participants provided written informed consent.

Due to the NeuroMET study design, in which follow-up assessments included only memory testing, as well as technical issues or contraindications for MRS (e.g., ferromagnetic implants), some follow-up visits in the current dataset lacked measurements of plasma p-tau181 (n = 60) or MRS-derived glutamate and GABA (n = 69). One participant in the CN Aβ+ group had missing MRS data at visit 1 but was retained in the analysis because MRS data were available at a subsequent visit. Group-specific missing data per visit and the intervals between follow-up visits are reported in Table 1.

**Table 1:** Observations grouped by diagnosis and visit. Time from visit 1 is presented as mean (SD). ‘n missing MRS’ indicates how many participants had no MRS data at that visit. Further statistical models might contain fewer observations according to data availability. Abbreviations: AD = Alzheimer’s Disease, Aβ+ = Amyloid-beta positive by plasma, CN = Cognitively Normal, MCI = Mild Cognitive Impairment, MRS = Magnetic Resonance Spectroscopy, V1 = Visit 1

| | CN Aβ- | CN Aβ+ | MCI Aβ+ | AD Aβ+ |
| --- | --- | --- | --- | --- |
| **Visit 1** |  |  |  |  |
| n | 43 | 17 | 20 | 24 |
| n missing MRS | 0 | 1 | 0 | 0 |
| time from V1 [years] | 0 (0) | 0 (0) | 0 (0) | 0 (0) |
| **Visit 2** |  |  |  |  |
| n | 38 | 14 | 15 | 18 |
| n missing MRS | 25 | 10 | 11 | 16 |
| time from V1 [years] | 1 (0.1) | 1 (0.1) | 1 (0.1) | 1.1 (0.3) |
| **Visit 3** |  |  |  |  |
| n | 20 | 9 | 4 | 6 |
| n missing MRS | 0 | 0 | 1 | 1 |
| time from V1 [years] | 3 (0.3) | 3.1 (0.2) | 3.1 (0.2) | 2.8 (0.5) |
| **Visit 4** |  |  |  |  |
| n | 15 | 9 | 2 | 2 |
| n missing MRS | 2 | 1 | 1 | 0 |
| time from V1 [years] | 4.1 (0.2) | 4.1 (0.3) | 4.1 (0.3) | 3.8 (0.2) |
| **Visit 5** |  |  |  |  |
| n | 6 | 2 | 0 | 0 |
| n missing MRS | 0 | 0 | 0 | 0 |
| time from V1 [years] | 5.2 (0.1) | 5.2 (0.1) |  |  |

### 2.2 AD-related Plasma Biomarkers

Plasma concentrations of p-tau181, Aβ42, Aβ40, NfL, and GFAP were quantified on the Simoa HD-X platform (4× onboard dilution) using the commercially available NEUROLOGY 4-PLEX E kit (Aβ40, Aβ42, GFAP, NfL; #103670) and the p-tau181 Advantage Kit V2 (#103714) at the Neurochemistry Lab, Amsterdam. Measurements of Aβ40, Aβ42, GFAP, and NfL were performed in singlicate, whereas p-tau181 was measured in duplicate, yielding a mean coefficient of variation of 5.1% (range 0.1–19.2%). Inter-assay quality was monitored using two in-house plasma pools, and all samples were analyzed in a single batch to ensure longitudinal consistency. Participants were fasting before blood collection. The p-tau181 threshold indicating amyloid pathology was established previously by the Neurochemistry Laboratory at Amsterdam UMC (Verberk et al., 2024). For the current study, the threshold value was adjusted to the specific kit lot by re-measuring 10 bridging samples. Based on this adapted threshold, amyloid status was defined as plasma p-tau181 > 2.08 pg/mL (Aβ+) or ≤ 2.08 pg/mL (Aβ-) at visit 1. To avoid misclassification due to measurement fluctuations around the threshold, participants were also classified Aβ+ when their plasma p-tau181 levels exceeded 2.02 pg/mL at visit 1 and 2.08 pg/mL at subsequent visits (n = 2). Apolipoprotein E (APOE) genotypes (rs429358 and rs7412) were determined using TaqMan assays, following established protocols (O’Dwyer et al., 2012).

### 2.3 Structural Magnetic Resonance Imaging and Analysis

High-resolution structural brain images were acquired on a 7T whole-body system (Magnetom 7T, Siemens Healthineers) using a 1Tx/32Rx-channel head coil. We used a 3D T1-weighted MP2RAGE sequence (Marques et al., 2010) with a denoised reconstruction approach (O’Brien et al., 2014). Imaging parameters were: TR/TE = 5000/2.51 ms, inversion times TI1/TI2 = 900/2700 ms, bandwidth 250 Hz/pixel, flip angles 7°/5°, isotropic voxel size 0.75 mm, and a field of view of 240 × 240 × 180 mm^3^. generalized auto-calibrating partial parallel acquisition (GRAPPA, acceleration factor 2, 32 reference lines). To quantify PCC + precuneus volumes as a covariate to account for neurodegeneration in one of our statistical models, we used the open-source FreeSurfer 7.1.1 image analysis suite (Martinos Center for Biomedical Imaging, Massachusetts General Hospital, http://surfer.nmr.mgh.harvard.edu/, Fischl (2012)). The full (pre-)processing pipeline and code were published earlier (Göschel et al., 2023).

### 2.4 Magnetic Resonance Spectroscopy Acquisition and Analysis

The MRS data were acquired using the same 7T scanner and head coil. Based on the structural image, a 20 × 20 × 20 mm^3^ voxel of interest (VOI) was positioned in the posterior cingulate cortex (PCC)/precuneus region, a metabolically active hub region (Leech & Sharp, 2014), as shown in the inlet of Figure 1. Localized RF calibration and second-order B0 shimming (Fillmer et al., 2015; Nassirpour et al., 2018) were performed prior to acquisition. Single-voxel spectra were collected with the spin echo full intensity acquired localization (SPECIAL) sequence (Mekle et al., 2009), including interleaved outer-volume suppression and variable power and optimized relaxation delays (VAPOR, Tkáč et al., 1999) water suppression (TR/TE = 6500/9 ms, 64 acquisitions, 2048 sample points, 512 ms acquisition time, 4000 Hz excitation bandwidth, 60 Hz water suppression bandwidth). In addition, a non–water-suppressed reference spectrum was acquired with otherwise identical parameters (4 acquisitions).

**Figure 1:**
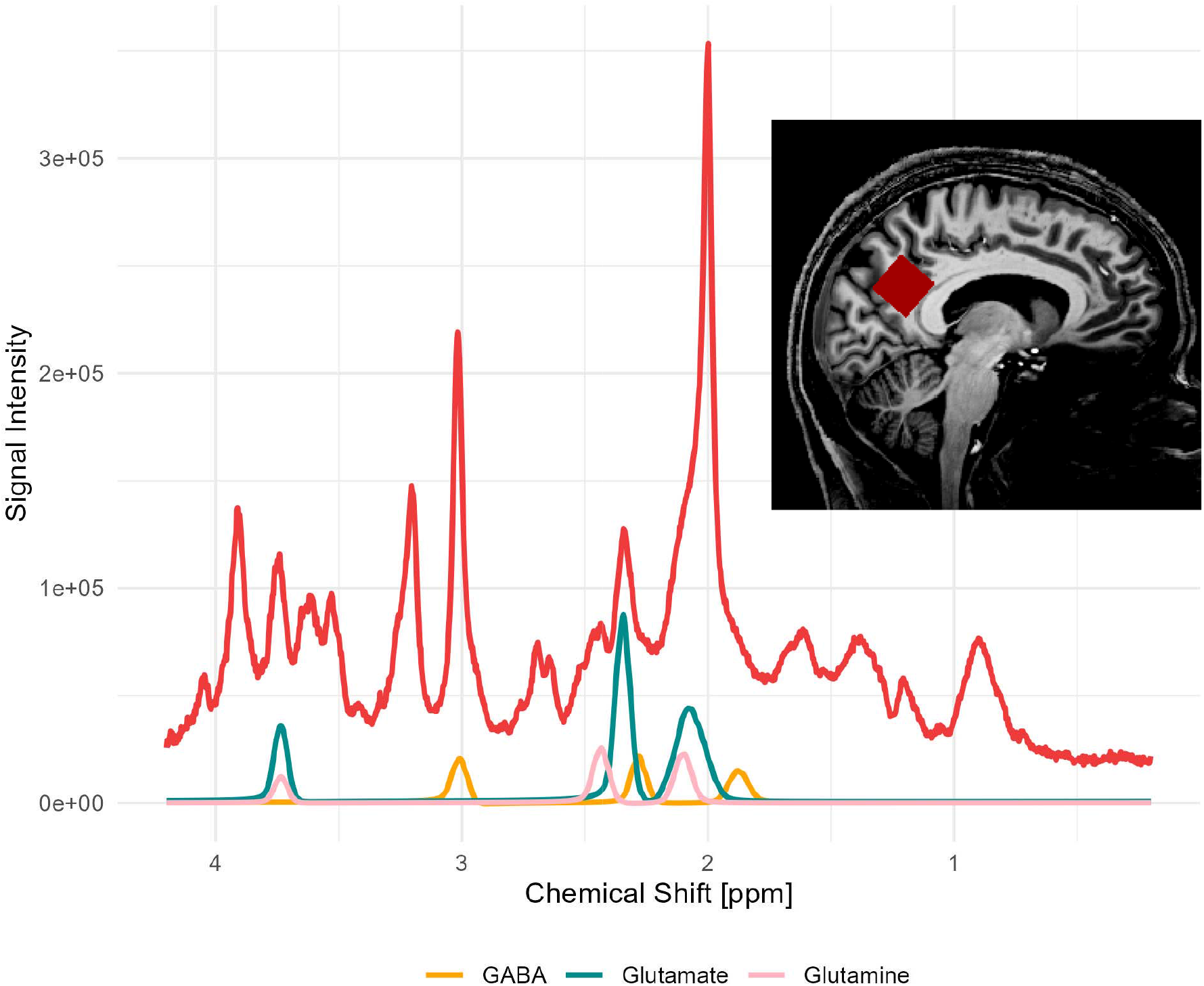
A 7T 1H-MRS spectrum acquired in the PCC/precuneus voxel (red square) from a participant of this study. (in their 70’s, subjective cognitive complaints, Aβ+). Due to spectral overlap, accurate quantification requires integrating multiple peaks. Glutamate is characterized by three multiplets at 2.04r⍰ppm, at 2.35r⍰ppm, and at 3.74r⍰ppm. GABA is characterized by three multiplets at 1.89, 2.28, and 3.01 ppm. Glutamine is characterized by three multiplets at 2.12, 2.45, and 3.75r⍰ppm. Abbreviations: GABA = gamma-aminobutyric acid, MRS = magnetic resonance spectroscopy, PCC = posterior cingulate cortex.

The MRS raw data were preprocessed using an in-house MATLAB pipeline previously described (Dell’Orco et al., 2024; Riemann et al., 2022). At first, each odd acquisition was averaged with the next even acquisition to yield fully localized spectra. A weighted, phase-corrected coil combination was applied, followed by frequency alignment to the NAA methyl resonance at 2.00 ppm. Before averaging, each localized signal was visually inspected, and motion-corrupted signals were discarded. Quantification was performed using LCModel v6.352 (Provencher, 2001) with a simulated basis set generated in VeSPA v0.9.3.53 (Soher et al., 2023) in combination with a single-component macromolecule as described earlier (Dell’Orco et al., 2024; Riemann et al., 2022). Signal intensities were water-scaled and corrected for tissue composition by segmenting the corresponding structural images into gray matter, white matter, and CSF (Near et al., 2021) to derive absolute metabolite concentrations for glutamate and GABA. Cramér–Rao lower bounds (CRLBs, Cavassila et al., 2001) were extracted from the LCModel output to provide an index of individual measurement reliability. Glutamate/GABA ratios were natural-log transformed to normalize the distribution, which was necessary because GABA has naturally lower concentrations in the brain than glutamate.

### 2.5 Functional Magnetic Resonance Imaging Acquisition at Resting-State and Analysis

Resting-state blood oxygen level–dependent (BOLD) functional MRI (fMRI) data were acquired with a multiband, multislice gradient-echo echo–echo planar imaging (EPI) sequence (Cauley et al., 2014; Moeller et al., 2010, TR = 2200 ms, TE = 28.2 ms, echo spacing = 0.97 ms, voxel size = 1.5 mm isotropic, FoV = 240 × 240 × 120 mm^3^, flip angle = 66°, GRAPPA acceleration factor = 2, 32 reference lines, multiband factor = 4, 300 volumes). Axial slices were aligned to the line connecting the anterior and posterior commissures. For distortion correction, an additional B0 field map was obtained with identical shim settings and orientation (TR = 500 ms, TE1/TE2 = 5/7.02 ms, 3 mm isotropic resolution). Participants were instructed to remain still, keep their eyes closed, and allow their thoughts to flow naturally. Preprocessing was carried out using fMRIPrep v20.1.1 (Esteban et al., 2019) and the fmriprep confounds used for denoising with NiLearn (Abraham et al., 2014), i.e., motion parameters, anatomical CompCor components, scrubbing regressors, and a high-pass filter. Subject-specific MRS voxel masks were coregistered to the MNI152NLin2009cAsym template, and seed time series were extracted using a standardized workflow comprising 6-mm FWHM spatial smoothing, z-scoring, and band-pass filtering (0.01–0.1 Hz). To obtain the whole-brain seed-to-voxel functional connectivities (FC), with the individual MRS voxel as seeds, masks of the MRS voxels were generated in NIfTI format from the voxel coordinates in the RDA files. FCs were then computed as Pearson correlations between the seed signal and each voxel’s time series. The full code was published on GitHub (Dell’Orco, 2026).

### 2.6 Memory Ability

Memory ability was assessed using the NeuroMET Memory Metric (NMM), a recently developed composite metric derived within the NeuroMET projects (Melin, Cano, Gillman, et al., 2023). The metric is based on a carefully selected item bank containing 57 dichotomous items, ranging from language- and culture-independent tasks (e.g., block recall, digit recall) to word-based recall items. Previous studies have shown that the NMM provides greater accuracy than conventional memory metrics, without compromising validity (Melin, Cano, Flöel, et al., 2023; Melin, Cano, Gillman, et al., 2023). We used years of education to adjust for the effects of cognitive reserve in statistical models, including the NMM.

### 2.7 Statistics

The statistical analyses for the main analysis were performed in R (version 4.5.1, R Core Team, 2025). The main models in this study were fitted using the lme4 package (Bates et al., 2025), and estimated marginal means and contrasts were obtained with the emmeans package (Lenth et al., 2025). Model visualizations were created using ggplot2 (Wickham, 2016). The full reproducible code was published on GitHub (Göschel, 2026).

Longitudinal associations between neurotransmitter concentrations, pathology markers, and memory ability were analyzed using linear mixed-effects models fitted with REML estimation and random intercepts for participants to account for within-participant dependence. Continuous predictors were modeled using natural splines (ns) to capture potential non-linear effects flexibly. Spline degrees of freedom (listed in Table A1) were chosen by comparison of AICs with alternative model parameterizations. Metabolite models (glutamate, GABA, glutamine) but not ratio models were weighted by CRLBs to account for measurement uncertainty. Four model families were specified:

#### Model 1

We tested whether longitudinal trajectories of neurotransmitter levels differed by diagnostic group, adjusting for year, age at visit 1, and sex:

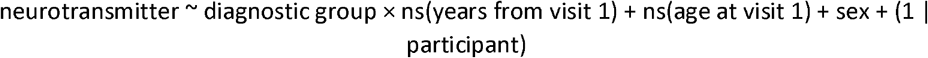

#### Model 2

We examined neurotransmitter levels as a function of continuous plasma p-Tau181, independently of diagnostic group, adjusting for year, age at visit 1, and sex:

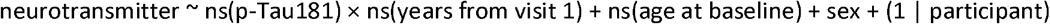

#### Model 3

We assessed whether memory ability (NMM) was associated with neurotransmitter levels across diagnostic groups, with adjustment for year, age at visit 1, sex, and education:

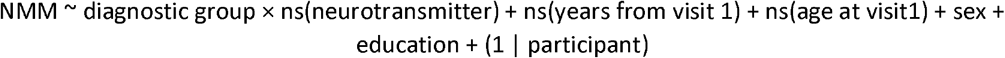

#### Model 4

We evaluated the relationship between astrocytic dysfunction and neurotransmitter measures involved in the glutamate-glutamine metabolism. Hence, we modeled glutamate, GABA, log(glutamate/GABA), glutamine, and log(glutamate/glutamine) as a function of plasma GFAP, adjusting for individual PCC + precuneus volume to account for neurodegeneration, year, age at visit 1, and sex:

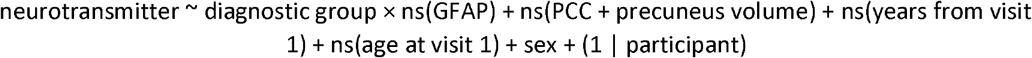

To assess the robustness of our longitudinal models, we performed sensitivity analyses excluding datapoints with disproportionate influence on model fit. One AD Aβ+ participant showed atypical glutamate, GABA, and glutamine values at visit 4, which disproportionately affected the group trajectory and led to a less robust nonlinear fit. As we were unable to identify any technical error or clinical explanation (Figure A1), this data point was retained in the dataset. However, because it obscured the otherwise consistent group-level patterns, we report the main trajectories in Figures 4B and 5 without this data point and provide results including this data point in the supplement (Figure A2).

For an additional proof-of-concept analysis, we assessed whether neurotransmitter levels were associated with resting-state seed-to-voxel FC. This analysis was carried out using Python and analogous modelled as a linear mixed effect model:

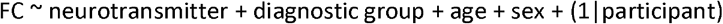

## 3. Results

### 3.1 Characteristics of the Study Population

Demographic, genetic, cognitive, metabolic, and biomarker data of all participants at visit 1 are summarized in Table 2. Groups differed in age (older in Aβ+ groups), sex distribution (fewer females in Aβ+ groups), and APOE ε4 carrier status (more frequent in MCI Aβ+ and AD Aβ+) (all p < 0.001). Cognitive performance, i.e., MMSE scores and memory ability, declined progressively from CN to AD. At the same time, plasma p-Tau181 and GFAP concentrations were higher in Aβ+ groups and increased with disease severity (all p < 0.001). In contrast, no substantial group differences were observed for education. As shown in Extended Figure 1, longitudinal plasma p-Tau181 increased in the CN Aβ- and AD Aβ+ groups, memory ability decreased in the AD Aβ+ group, and plasma GFAP increased in the CN Aβ-, CN Aβ+, and AD Aβ+ groups.

**Table 2:** Descriptive statistics and group comparisons of the characteristics of the study population at visit 1. For continuous variables (e.g., age, MMSE, metabolite concentrations), the mean and 95% confidence interval are shown. P-values are derived from linear models: for MRS data and plasma p-Tau181, the models are adjusted for age and sex; for MMSE and memory (NMM), the models are additionally adjusted for education. For categorical variables (sex and APOEε4 status), absolute and relative frequencies are presented, and p-values are based on Chi-square tests. Abbreviations: AD = Alzheimer’s Disease, APOE ε4 = Apolipoprotein E epsilon 4 allele, Aβ+ = Amyloid-beta positive by plasma, CN = Cognitively Normal, GABA = Gamma-Aminobutyric Acid, MCI = Mild Cognitive Impairment, MMSE = Mini-Mental State Examination, MRS = Magnetic Resonance Spectroscopy, p-Tau181 = phosphorylated tau at threonine 181.

| Variable | CN Aβ- | CN Aβ+ | MCI Aβ+ | AD Aβ+ | p-value |
| --- | --- | --- | --- | --- | --- |
| Sample size | n = 43 | n = 17 | n = 20 | n = 24 | NA |
| Age [years] | 67.9 [65.9;70.0] | 74.9 [71.4;78.3] | 72.8 [70.3;75.4] | 75.9 [73.3;78.5] | <0.001 |
| Sex (Female) | 29 (67%) | 6 (35%) | 7 (35%) | 12 (50%) | <0.001 |
| Education [years] | 15.0 [14.2;15.8] | 16.1 [14.4;17.8] | 15.0 [13.6;16.4] | 14.1 [13.0;15.2] | 0.184 |
| APOE ε4 (carrier) | 12 (28%) | 3 (18%) | 13 (65%) | 15 (62%) | <0.001 |
| MMSE | 29.1 [28.8;29.4] | 29.1 [28.6;29.5] | 26.9 [25.9;27.8] | 22.6 [20.6;24.7] | <0.001 |
| Memory Ability (NMM) [logits] | 0.69 [0.48;0.91] | 0.61 [0.31;0.91] | -0.33 [-0.61;-0.05] | -1.26 [-1.62;-0.90] | <0.001 |
| MRS Glutamate [mmol/L] | 13.2 [12.8;13.6] | 13.5 [12.7;14.3] | 12.6 [12.1;13.2] | 13.2 [12.5;13.9] | 0.278 |
| MRS GABA [mmol/L] | 3.51 [3.33;3.69] | 3.65 [3.38;3.91] | 3.39 [3.09;3.69] | 3.48 [3.21;3.74] | 0.618 |
| log(Glutamate/GABA) | 1.33 [1.28;1.39] | 1.31 [1.25;1.37] | 1.33 [1.25;1.42] | 1.35 [1.27;1.43] | 0.933 |
| Plasma p-Tau181 [pg/mL] | 1.45 [1.35;1.55] | 2.94 [2.50;3.38] | 3.17 [2.52;3.82] | 3.93 [3.08;4.79] | <0.001 |
| Plasma GFAP [pg/mL] | 100.5 [89.0;111.9] | 137.8 [107.9;167.6] | 164.7 [126.8;202.6] | 236.0 [188.9;283.2] | <0.001 |

### 3.2 Glutamate and GABA show stage-dependent alterations

To test whether glutamate and GABA follow non-linear trajectories across AD progression, we modeled longitudinal neurotransmitter levels as a function of diagnostic group and time (years from visit 1) using linear mixed-effects models, adjusting for age at baseline and sex (model 1). At visit 1, levels of 7T MRS glutamate, GABA, and log(glutamate/GABA) did not differ substantially between diagnostic groups (all p ≥ 0.104 for glutamate, p ≥ 0.541 for GABA, p ≥ 0.911 for log ratio; Figure 2A; mean values in Table 2). Longitudinally, however, group-specific changes emerged (Figure 2B): Glutamate levels remained stable in CN Aβ–, increased slightly in CN Aβ+ (0.20 pg/ml per year) and more strongly in MCI Aβ+ (0.80 pg/ml per year), but decreased in AD Aβ+ (–0.44 pg/ml per year). More moderately, GABA levels followed the increases in glutamate during the pre-dementia stages CN Aβ+ and MCI Aβ+ (0.12 and 0.17 pg/ml per year, respectively). In contrast to glutamate, GABA remained stable in the AD Aβ+ group (-0.01 pg/ml per year). Given that glutamate is more abundant (mean for CN Aβ– = 13.2 pg/ml) than GABA (mean in CN Aβ– = 3.5 pg/ml), these small absolute changes in GABA represent proportionally similar magnitudes to glutamate. Correspondingly, the rising GABA levels appeared to balance glutamate-dependent changes, resulting in stable log(glutamate/GABA) ratios in predementia stages. However, the log(glutamate/GABA) ratio showed a slight decrease in CN Aβ– (–0.02 per year) and AD Aβ+ (-0.04 per year, trend significant).

**Figure 2:**
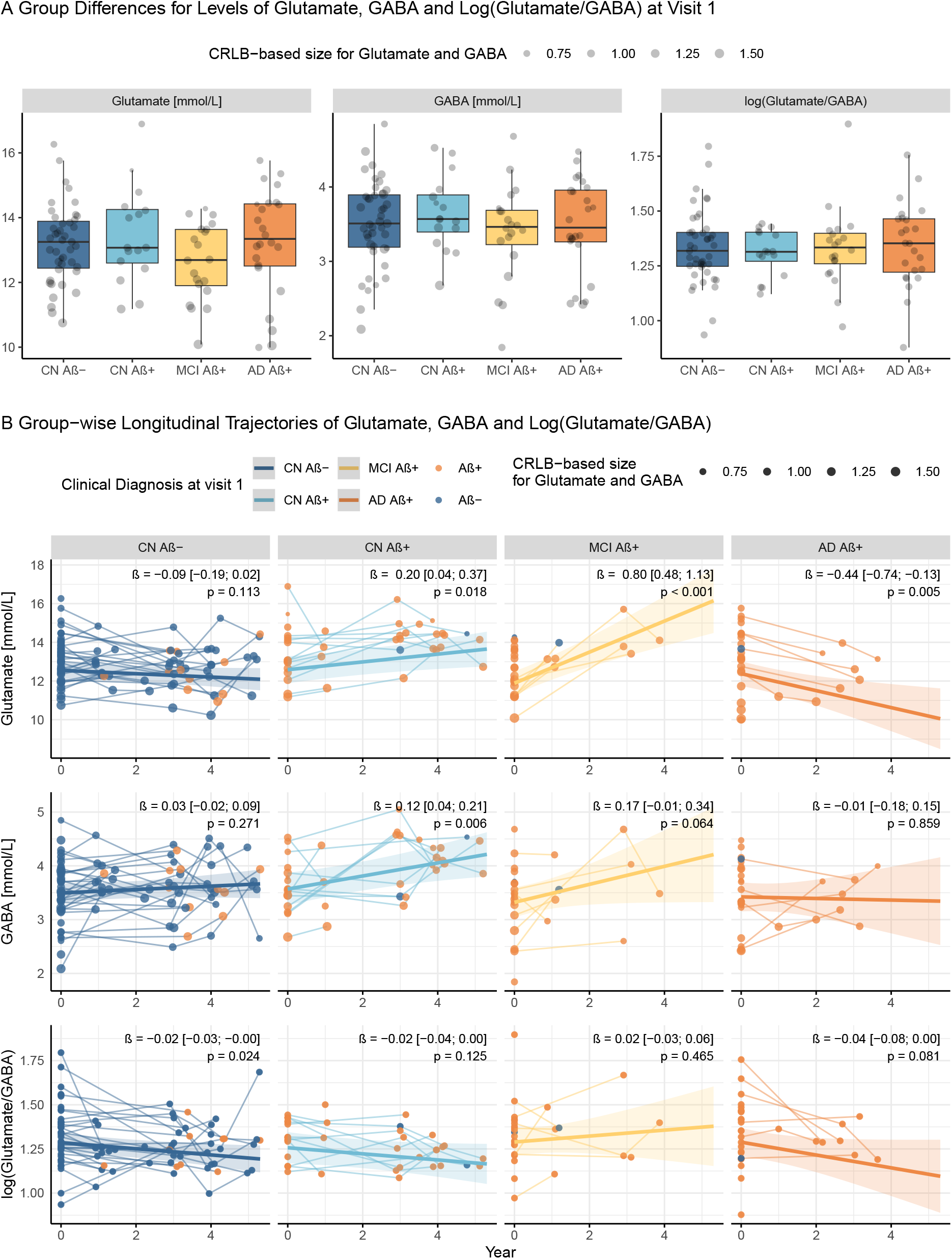
Group comparisons of neurotransmitter concentrations measured by 7T MRS at visit 1 (A) and their longitudinal changes (B). (A) At visit 1, concentrations of MRS glutamate, GABA, and log(glutamate/GABA) did not differ substantially across diagnostic groups. (B) Model 1 showed that longitudinal glutamate levels increased in the CN Aβ+ and MCI Aβ+ groups, and decreased in the AD Aβ+ group. GABA levels increased in the CN Aβ+ and MCI Aβ+ groups. In contrast, the glutamate/GABA ratio declined significantly in the CN Aβ-, and AD Aβ+ groups. Displayed β coefficients and p-values indicate estimated annual change per group, based on estimated marginal trends of model 1. Datapoints were sized using Cramér–Rao lower bounds (CRLB), a common metric for quantifying uncertainty in MRS. Here, smaller datapoints represent higher uncertainty (i.e., higher CLRBs). Amyloid β positivity (Aβ +) was defined as plasma p-Tau181 > 2.08 pg/mL, as indicated by the orange points and dashed line. Abbreviations: AD = Alzheimer’s disease, Aβ = amyloid-beta, CN = cognitively normal, CRLB = Cramér-Rao lower bound, GABA = gamma-aminobutyric acid, MCI = mild cognitive impairment, MRS = magnetic resonance spectroscopy, p-Tau181 = tau phosphorylated at threonine 181.

Sensitivity analyses that retained one disproportionately influential datapoint in the AD Aβ+ group suggested a more complex, U-shaped trajectory for glutamate, especially in AD Aβ+ (Supplementary Figure A2).

Together, these findings demonstrate stage-dependent alterations in excitatory and inhibitory neurotransmission across the AD continuum, characterized by increasing glutamate levels in amyloid-positive pre-dementia stages and a subsequent decline in AD dementia, whereas changes in GABA or the GABA ratio were more moderate.

### 3.3 Plasma p-Tau181 alone was not a substantial Modulator of the observed Changes of Glutamate or GABA

To determine whether the observed stage-dependent trajectories of glutamate and GABA are better explained by continuous pathologic progression rather than cognition-based staging, we examined neurotransmitter levels as a function of longitudinal plasma p-Tau181 and time (years from visit 1), adjusting for age at visit 1 and sex (model 2). There were no substantial linear or non-linear associations between the continuous measure of plasma p-Tau181 and glutamate, GABA, or the glutamate/GABA ratio (Figure 3 A). Since the diagnostic group was derived from plasma p-Tau181 levels, we did not explore any p-Tau181 × diagnosis interaction terms. In line with the hypothesis of a biphasic trajectory of glutamate, however, supplementary model A1, comprising three-way interactions and thus with limited power, suggested that steeper longitudinal increases in plasma p-Tau181 slopes (presumably further advanced patients) might be associated with slower increases in glutamate in MCI Aβ+ individuals (Figure A3). In summary, the observed longitudinal group-wise changes in glutamate and GABA were better predicted by diagnostic groups (i.e., cognition-based staging) than by continuous plasma p-Tau181.

**Figure 3:**
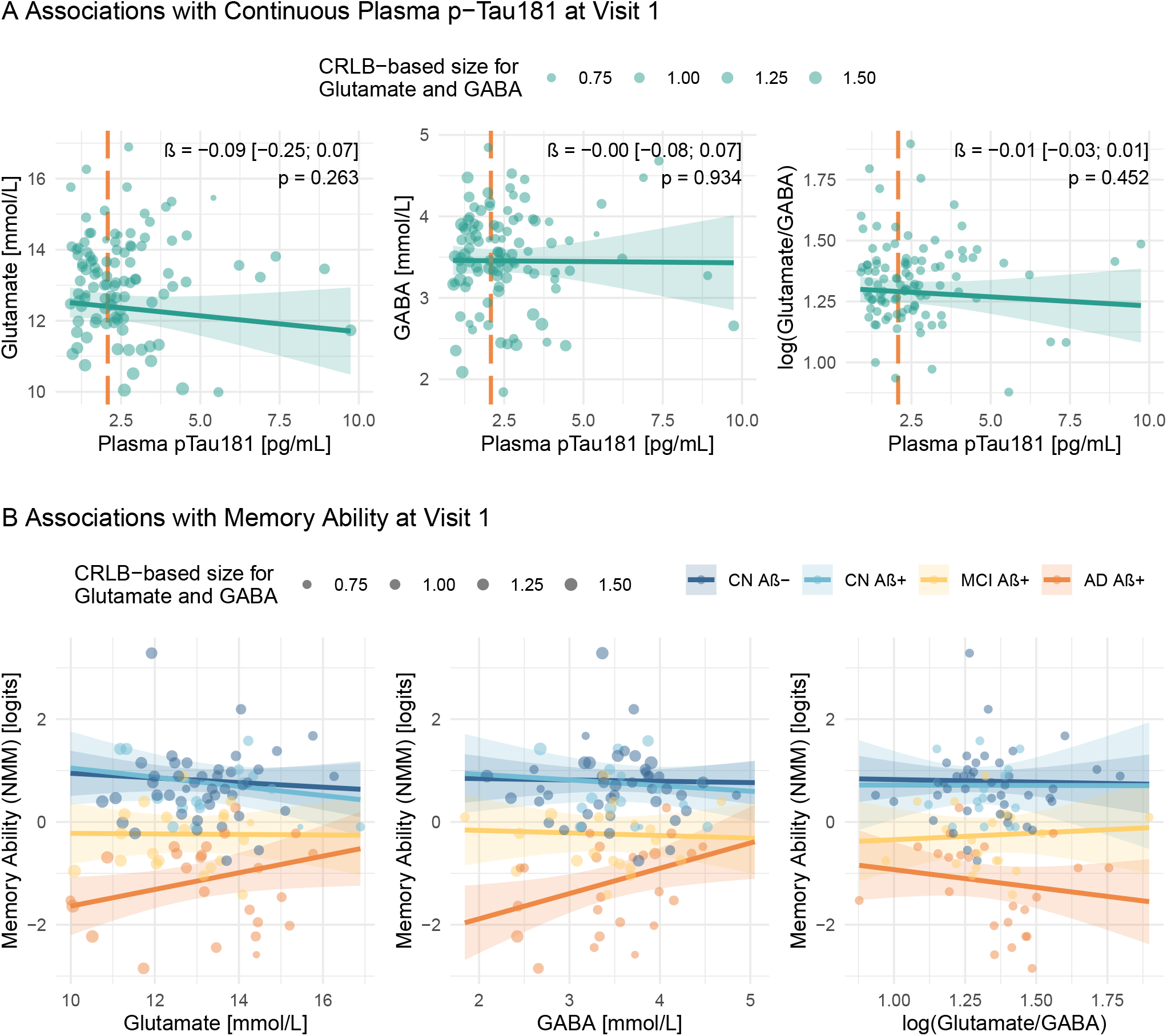
Visit 1 associations between 7T MRS glutamate, GABA, and log(glutamate/GABA) concentrations with (A) plasma p-Tau 181 and (B) memory ability measured by NMM. (A) No substantial associations were observed with the continuous measure of plasma p-Tau181 (independent of diagnostic group). (B) Higher glutamate and GABA concentrations were associated with better memory ability only in the AD Aβ+ group. For (A), fits were derived from linear mixed-effects models adjusted for age and sex (Model 2), and for (B), additionally for diagnostic group and education (Model 3). Shaded areas represent 95% confidence intervals. Individual data points at visit 1 are overlaid. Glutamate and GABA models were weighted by the Cramér–Rao lower bounds (CRLBs), with smaller points indicating higher uncertainty (i.e., larger CRLBs). The orange dashed line in (A) marks the plasma p-Tau181 threshold (>2.08 pg/mL) used to classify amyloid positivity (Aβ+). Abbreviations: AD = Alzheimer’s disease, Aβ = amyloid-beta, CN = cognitively normal, CRLB = Cramér-Rao lower bound, GABA = gamma-aminobutyric acid, MCI = mild cognitive impairment, MRS = magnetic resonance spectroscopy, NMM= NeuroMET Memory Metric, p-Tau 181 = tau phosphorylated at threonine 181.

### 3.4 Higher glutamate and GABA concentrations were associated with better memory performance only in Aβ+ AD

To examine whether stage-dependent changes in glutamate and GABA are functionally relevant for cognition across diagnostic groups, we modelled memory ability (indicated by NMM) as a function of neurotransmitter concentration, diagnostic group, and time (years from visit 1), adjusting for age at visit 1, sex, and education (model 3). As shown in Figure 3B, substantial associations were observed only in the AD Aβ+ group: better memory ability was associated with both higher glutamate (β = 0.16 [0.00; 0.32], p = 0.046) and higher GABA levels (β = 0.49 [0.09; 0.90], p = 0.018). In contrast, the log(glutamate/GABA) ratio showed no significant association in the AD Aβ+ group (β = -0.69 [-2.04; 0.65], p = 0.311). No associations to memory ability were detected in individuals of the CN Aβ-, CN Aβ+, and MCI Aβ+ groups (all p ≥ 0.322). These findings suggest that, in predementia stages, when glutamate and GABA levels were observed to rise simultaneously (a homeostatic response), these levels did not predict memory functioning. Only in the dementia stage, when the glutamate and GABA trajectories diverge, are their levels associated with memory ability.

### 3.5 Astrocytic Dysfunction measured by plasma GFAP might impair the glutamate-glutamine cycle, but was not associated with elevated glutamate levels

We further investigated whether astrocytic reactivity, indicated by elevated plasma GFAP, is associated with the AD-related dynamic changes in glutamate and GABA. Therefore, we examined the group-wise association between plasma GFAP and 7T MRS neurotransmitters, including the glutamate-glutamine ratio, which is a surrogate for astrocytic glutamate-glutamine cycling. We adjusted for time (years from visit 1), age at visit 1, sex, and PCC+precuneus volume as an estimate of neurodegeneration progression within the MRS voxel region (model 4, Figure 4B). Post hoc analyses of the estimated marginal means revealed important effects: Overall, astrocytic dysfunction, as indicated by plasma GFAP, was not significantly associated with elevated glutamate and GABA levels in AD pathology. Only in the AD Aβ+ group, plasma GFAP was associated with a trend towards lower glutamate levels (β_Glutamate∼GFAP_ = -3.68 [-8.03; 0.67]) × 10^3^, p = 0.096). However, higher plasma GFAP levels were associated with lower glutamine in the AD Aβ+ (-2.66 [-4.65; -0.66]) × 10^3^, p = 0.009). A similar relation was observed at trend level in the CN Aβ-(β_Glutamine∼GFAP_ = -3.26 [-6.84; 0.32]) × 10^3^, p = 0.074) and MCI Aβ+ groups (-2.49 [-5.19; 0.21]) × 10^3^, p = 0.071), but not in the CN Aβ+ group (0.05 [-3.53; 3.63]) × 10^3^, p = 0.977). In the CN Aβ- and MCI Aβ+ groups, this lead to an association between higher plasma GFAP and increased ratios of glutamate and glutamine (CN Aβ-: β_log(Glutamate/Glutamine)∼GFAP_ = 0.79 [0.01; 1.57]) × 10^3^, p = 0.046, MCI Aβ+: β_log(Glutamate/Glutamine)∼GFAP_ = 0.50 [-0.09; 1.09]) × 10^3^, p = 0.096). In summary, we showed that although higher GFAP was associated with lower glutamine levels in both groups with and without amyloid pathology, it was not associated with the previously described increase in glutamate levels in early amyloid pathology.

**Figure 4:**
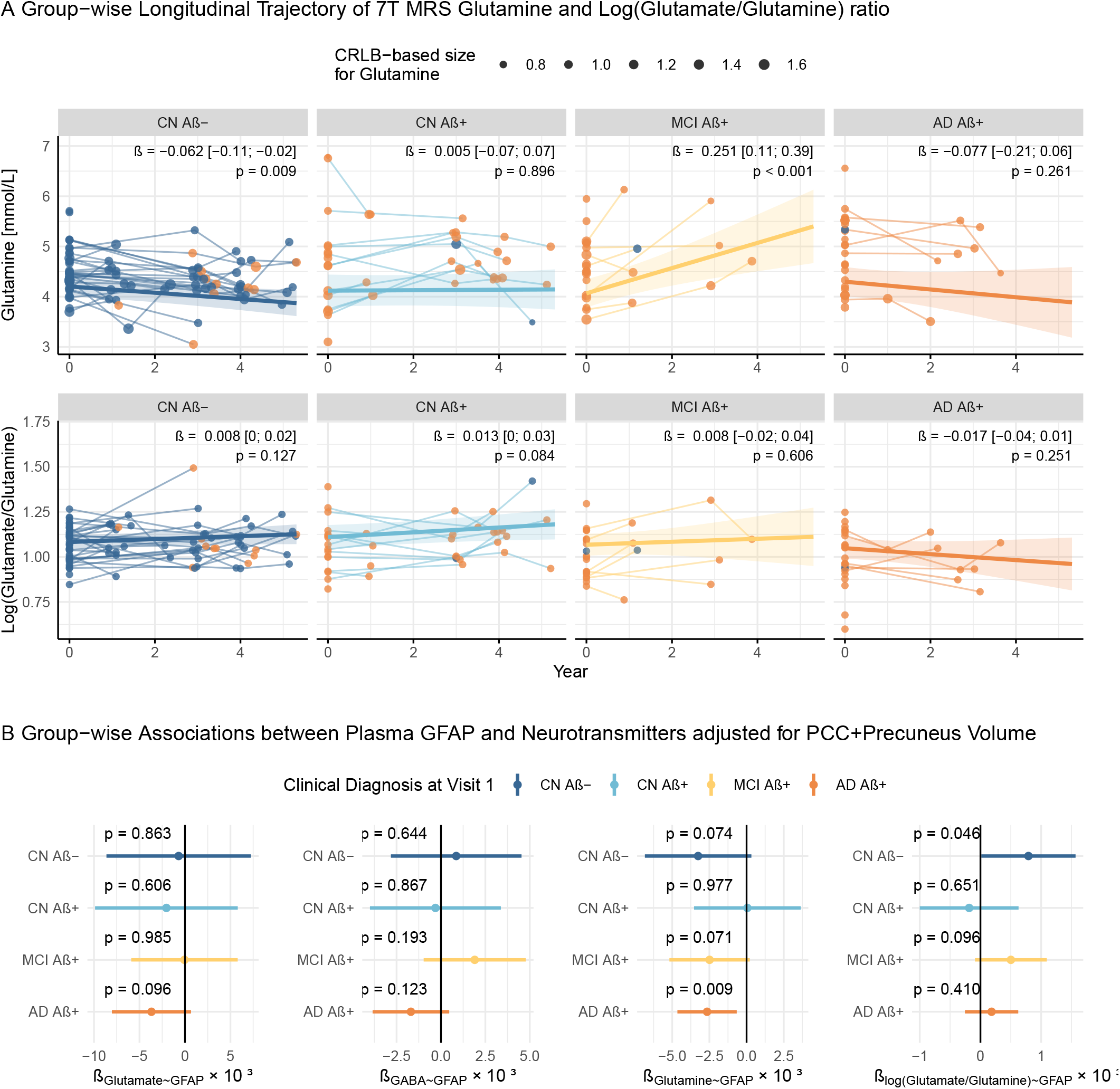
Reactive astrocytes, indicated by elevated GFAP, were associated with the glutamate-glutamine cycling. A) Longitudinal trajectories of glutamine and Log(glutamate/glutamine) levels by diagnostic group, derived from model 1 adjusted for age and sex. B) Associations between plasma GFAP and glutamate, GABA, glutamine, and the log(glutamate/glutamine) ratio measured by 7T MRS. Estimated marginal mean slopes suggested that higher plasma GFAP levels were not associated with higher glutamate levels, as previously observed in early amyloid pathology. However, there were group-specific associations and trends between higher GFAP levels and lower glutamine levels, lower glutamate levels, or higher log(glutamate/glutamine) ratios. These results were derived from linear mixed-effects models that included a diagnostic group × GFAP interaction (Model 4), adjusted for age, sex, year, and PCC+precuneus volume. Shaded areas and error bars represent the 95% confidence intervals. Except for the log(glutamate/glutamine) model, all models were weighted by Cramér–Rao lower bounds (CRLB). Abbreviations: AD = Alzheimer’s disease, Aβ = amyloid-beta, CN = cognitively normal, CRLB = Cramér-Rao lower bound, GABA = gamma-aminobutyric acid, MCI = mild cognitive impairment, MRS = magnetic resonance spectroscopy.

### 3.6 Glutamate and GABA correlate with resting-state seed-to-voxel connectivity

Confirming the validity of our results, glutamate, GABA, and the log(glutamate/GABA) ratio showed positive correlations with resting-state functional connectivity from the individual MRS voxel locations to multiple other brain regions (Figure 5). The linear mixed-effects model was adjusted for group, age, and sex. Only regions showing β > 3.1, corresponding to p < 0.001, are displayed in Figure 5. The spatial patterns of these relevant associations were largely overlapping across glutamate, GABA, and the ratio. However, the strength of associations varied, with glutamate showing the strongest effect. These results suggest that resting-state fMRI connectivity, as used in previous neuroimaging studies of hyperconnectivity, is related to neurotransmitter levels.

**Figure 5:**
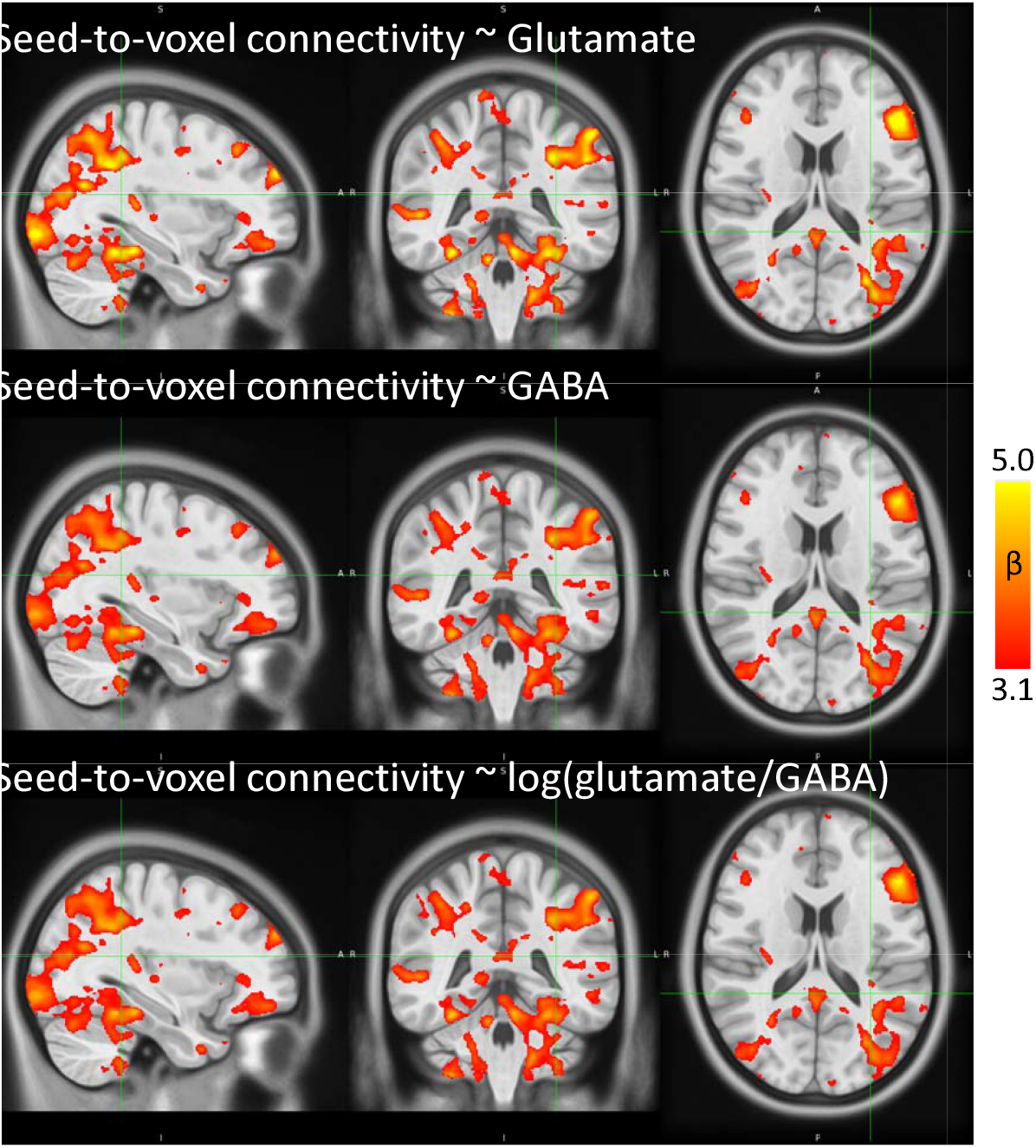
Brain regions showing significant positive associations between glutamate, GABA, and the log(glutamate/GABA) ratio and resting-state seed-to-voxel connectivity. Color scale reflects β coefficients ranging from 3.1 to 5 (red → yellow), corresponding to p < 0.001. The seed region was defined as the individual MRS voxel. Abbreviations: GABA = gamma-aminobutyric acid, MRS = magnetic resonance spectroscopy.

### 3.7 Lower levels of glutamate around age 70 and in female participants

It is worth noting that model 1 further uncovered that glutamate levels declined until about age 70 (first spline term β = -1.51, p = 0.066), after which they increased again (second spline term β = 0.58, p = 0.199), as shown in Extended Figure 2. The effect of age at visit 1 on GABA and log(glutamate/GABA) was not substantial (GABA: ß [95% CI] = -0.15 [-0.68; 0.42], p = 0.642; log(glutamate/GABA): F(2, 74.15) = 2.23, p = 0.115). Thus, glutamate levels may be more sensitive to age-related changes than GABA or their ratio. Furthermore, female participants showed significantly lower glutamate levels than males, with an estimated difference of -0.78 mmol/L (95% CI = [-1.24; - 0.33], p = < 0.001, Extended Figure 2D). In contrast, GABA levels did not differ between sexes; the estimated difference was -0.08 mmol/L (95% CI = [-0.29; 0.13], p = 0.431, Extended Figure 2). Similarly, no sex difference was observed in the log-transformed glutamate/GABA ratio (-0.04 mmol/L, 95% CI = [-0.09; 0.02], p = 0.186, Extended Figure 2).

## 4. Discussion

Our study addressed a critical gap in AD research by linking prior clinical neuroimaging studies of biphasic neuronal activity with animal-model studies suggesting glutamatergic involvement in hyperexcitation. By providing the first longitudinal characterization of cerebral glutamate and GABA dynamics in individuals across the AD continuum using high-field 7T MRS, we demonstrated that the trajectory of glutamate indeed suggests a hyperexcitation in early amyloid pathology, i.e., the CN Aβ+ and MCI Aβ+ groups, followed by a hypoexcitation in the later stage, i.e., the AD Aβ+ group. Dynamics of GABA showed modest longitudinal increases observed in the CN Aβ+ and MCI Aβ+ groups. Furthermore, we found in-vivo evidence that astrocytic dysfunction, indicated by higher plasma levels of GFAP, was involved in the deregulation of neurotransmitter concentrations: higher plasma levels of GFAP were associated with lower glutamine levels, achieving statistical significance in the AD Aβ+ group, and a higher glutamate/glutamine ratio in the CN Aβ-. Together, these data suggest glutamate-dependent mechanisms as a potential therapeutic target against AD but should account for the stage-dependent biphasic dynamics, alongside concurrent astrocytic metabolic failure.

### 4.1 The Biphasic Glutamate Trajectory

The key findings of this study are stage-dependent alterations in absolute glutamate levels, characterized by initial increases in the CN Aβ+ and MCI Aβ+ groups and a subsequent decline in the AD Aβ+ group. Together with the observation that higher glutamate levels were associated with stronger resting-state connectivity, our results provide further support for the proposed biphasic trajectory of neuronal activity in AD (Gallego-Rudolf et al., 2024). The observed increases in glutamate in the CN Aβ+ and MCI Aβ+ groups in this study reflect early hyperexcitability likely driven by Aβ pathology. Preclinical studies have shown that soluble Aβ oligomers, in particular, enhance glutamate release and alter receptor function (Busche et al., 2012; Hector & Brouillette, 2021; Rudy et al., 2015; Zott & Konnerth, 2023). The subsequent decrease in glutamate levels in the AD Aβ+ group is widely interpreted as a direct consequence of glutamate-mediated excitotoxicity, which leads to progressive neurodegeneration and synaptic loss (Satarker et al., 2022). Gallego-Rudolf et al. (2024) further suggested that the transition from hyperactivity to hypoactivity might correspond to the onset of tau pathology accumulation. Our results are mostly in contrast with the three 7T MRS studies of MCI or AD to date, which reported reduced glutamate/creatine ratios in MCI (Oeltzschner et al., 2019), unchanged glutamate in AD (Marjańska et al., 2019), and continuously decreasing glutamate in MCI and AD (Wong et al., 2020), considering that creatine levels were thought to be a “stable” internal reference. However, in line with our hypothesis, a 7T study of cognitively healthy individuals with amyloid pathology (corresponding to our CN Aß+ group) showed elevated glutamate/creatine ratios (Schreiner et al., 2024) and other MRS studies at lower field strengths observed lower glutamate or glutamate/creatine ratios in later stages of AD (Fayed et al., 2011; Rupsingh et al., 2011). Previous MRS studies were not able to capture the full non-linear trajectory, likely because of 1) the limited spectral resolution at lower field strengths, 2) the absence of longitudinal data, and 3) the lack of amyloid-based staging. Cross-sectional group differences were also not measurable in our data, which further emphasizes the importance of longitudinal study designs for characterizing disease progression. Overall, our in-vivo 7T MRS data provide a mechanistic bridge between human imaging studies showing biphasic trajectories of neuronal activity in AD using fMRI (Foster et al., 2018; Giorgio et al., 2024; Roemer-Cassiano et al., 2025) or MEG (Gallego-Rudolf et al., 2024), and animal-model studies suggesting non-linear glutamate-related pathophysiology (Rudy et al., 2015; Satarker et al., 2022; Zott et al., 2019; Zott & Konnerth, 2023).

### 4.2 A Transient Homeostatic Response Through Increased GABA That Ultimately Fails

Similar to glutamate, GABA levels increased in the CN Aβ+ and MCI Aβ+ groups, which may represent a homeostatic response to the Aβ-related hyperexcitability by glutamate (Palop et al., 2007). The stability of the log(glutamate/GABA) ratio in these groups likely reflects a balance between excitatory and inhibitory signaling regulated by the glutamate/GABA–glutamine cycle (Andersen, 2025). In a homeostatic system, GABAergic inhibition should closely follow glutamatergic excitation to prevent epileptiform overexcitation (Barbour et al., 2024) or cognitive impairment from excessive inhibition (Wu et al., 2014). Consequently, Marjańska et al. (2019), Oeltzschner et al. (2019), and Schreiner et al. (2024) showed GABA levels following glutamate levels in their cross-sectional 7T MRS studies. We further showed that GABA levels remained stable in the AD Aβ+ group, suggesting that the balancing mechanism ultimately fails, potentially due to the excitotoxic effects of excessive glutamate release, which can lead to neuronal death (Satarker et al., 2022). Unlike glutamate, GABA is not excitotoxic, and GABAergic neurons might be comparatively preserved during disease progression (Amorim et al., 2017), which is a possible explanation for the stability of GABA concentrations in the AD Aβ+ group of this study. Notably, only in the AD Aβ+ stage, when the log(glutamate/GABA) ratio shifted toward relatively lower glutamate levels, did both neurotransmitters correlate with memory ability. Our findings align with reports of abnormally high GABA levels in transgenic mouse models and human post-mortem tissue (Palop et al., 2007; Wu et al., 2014), but are in contrast with the widespread GABAergic deficits described in a recent review (Carello-Collar et al., 2023) and functional imaging studies showing a progressive shift from an excitatory–inhibitory homeostasis toward excitation (Li et al., 2025; Scaduto et al., 2023; van Nifterick et al., 2023). These inconsistencies in previous reports again highlight the importance of longitudinal, Aβ−integrating designs to capture the non-linear dynamics of excitatory–inhibitory balance. Together, our results support a model in which GABA transiently increases to stabilize network activity in early AD stages, but this homeostatic mechanism eventually collapses as neurodegeneration progresses.

### 4.3 Astrocytic Dysfunction and Impaired Glutamate–Glutamine Cycling

Higher plasma GFAP levels were associated with lower glutamine levels in the CN Aβ−, MCI Aβ+, and AD Aβ+ groups, and higher log(glutamate/glutamine) ratios in the CN Aβ− and MCI Aβ+ groups. Astrocytes play a central role in maintaining excitatory–inhibitory homeostasis by clearing extracellular glutamate and converting it into glutamine, which neurons recycle into glutamate and GABA (Rudy et al., 2015; Targa Dias Anastacio et al., 2022). Previous studies have shown that reactive astrocytes in AD exhibit impaired glutamate uptake and reduced glutamine synthetase expression (Targa Dias Anastacio et al., 2022; Zott & Konnerth, 2023), thereby compromising glutamate– glutamine homeostasis. This could explain the observed reduction in glutamine and the increase in the log(glutamate/glutamine) ratio with increasing astrocytic reactivity, as indicated by elevated plasma GFAP. The inclusion of the CN Aβ− group in this relationship suggests that the effect of reactive astrocytes on glutamine metabolism may be independent of amyloid pathology. Notably, no substantial associations between plasma GFAP and glutamate or GABA were observed, except for a trend towards lower glutamate in the AD Aβ+ group. As the disease progresses, advanced neurodegeneration and simultaneously reduced glutamate pools may mask the metabolic disturbances, despite correction for PCC+precuneus volume. Although several of these reported associations were only at the trend level, their directionality supports the interpretation that astrocytic dysfunction contributes to early AD-related metabolic changes, primarily through impaired glutamate-glutamine cycling. Overall, our results suggest that reactive astrocytes, reflected by elevated plasma GFAP, contribute to a malfunction of the glutamate-glutamine cycle early in the disease.

### 4.4 Clinical and Translational Implications

Overall, our findings support the hypothesis that AD is not a static process of neuronal loss but a dynamic alteration in excitation and inhibition over time. A recent study showed that hyperconnectivity accelerates tau propagation (Roemer-Cassiano et al., 2025). Modulating hyperexcitability could slow the spread of tau pathology and, in turn, clinical progression. Therapies tailored to these evolving phases, rather than uniform approaches across all stages, could substantially improve the effectiveness of disease-modifying interventions. Interventions in early stages (here, CN Aβ+ and MCI Aβ+ stages) could target glutamate-mediated hyperactivity, thereby preventing excitotoxic cell death and slow tau spread. Potential approaches have been suggested earlier, including modulation of glutamate release, enhancement of astrocytic glutamate uptake (e.g., via ceftriaxone), or use of antiepileptic or stabilizing agents such as levetiracetam and rapamycin (Barbour et al., 2024; Hector & Brouillette, 2021; Zott & Konnerth, 2023). Once neuronal activity declines (here, in the AD Aβ+ stage), anti-excitotoxic treatments may, however, become counterproductive. Together with previous literature, our findings highlight the importance of stage-specific therapeutic strategies for personalized medicine in AD.

### 4.5 Limitations

There are several limitations to consider when interpreting this study. First, the relatively small sample size limited the statistical power of the analyses. Especially in the MCI and AD groups, longitudinal data were incomplete for several participants. In particular, one influential data point substantially affected the longitudinal results and therefore had to be removed from the main analyses (the corresponding results, including the data point, are reported in the Supplementary Material). While the primary effects appeared robust, the absence of significant group interactions in the GFAP analyses may reflect limited power. Second, the dataset lacked confirmation of Aβ status using CSF or PET biomarkers for some participants. Consequently, plasma p-Tau181 was used as a proxy to estimate Aβ positivity. While this approach has been validated and correlates well with established biomarkers (Verberk et al., 2024), it remains less precise than CSF or PET measures. To mitigate this limitation, we were transparent about Aβ status throughout the analyses and visualized it by color-coding data points in all relevant figures. Third, there are technical and methodological limitations inherent to 7T MRS. Despite its superior spectral resolution and signal-to-noise ratio compared to lower field strengths, 7T MRS remains sensitive to head motion, leading to increased magnetic field inhomogeneity. Additionally, the limited voxel size and placement variability may affect between-subject comparability and it is not possible to differentiate between intra- and extracellular neurotransmitter concentrations. Despite these limitations, this study combines, for the first time, longitudinal 7T MRS, functional imaging, plasma biomarkers, and memory assessments to explore neurochemical alterations across AD stages, a combination rarely achieved in human studies.

### 4.6 Conclusion

This study provides the first longitudinal characterization of cerebral glutamate and GABA dynamics across the AD continuum using ultra–high-field 7T MRS. Our findings built a mechanistic bridge between previous neuroimaging studies of hyperactivity/-connectivity in humans and studies of amyloid-induced hyperexcitation with increased glutamate in mouse models. Our findings outline an early phase of glutamate-driven hyperexcitation during pre-dementia stages, followed by a later phase of hypoexcitation as neurodegeneration progresses. Concurrently, GABA showed modest early increases, potentially reflecting a transient compensatory response that ultimately fails. The observed associations between astrocytic dysfunction, as indicated by plasma GFAP, and lower glutamine levels further suggest that an impaired glutamate–glutamine cycling might be an essential mediator linking amyloid pathology to hyperexcitation. Together with the discussed literature, our findings redefine AD progression as a dynamic interplay between neuronal excitability and astrocytic metabolism, rather than a linear decline in neuronal function. Modulating hyperexcitability may slow downstream spread of tau pathology and thus clinical progression, a hypothesis to be tested in future clinical trials.

## Data Availability

All data produced in the present study are available upon reasonable request to the authors

https://doi.org/10.5281/ZENODO.18254795

https://doi.org/10.5281/zenodo.18266073

## Acknowledgements

The authors thank the NeuroMET and NeuroMET2 consortia for their cooperation, Drs. Jens Bohlken and Sonja Fabian for their efforts in recruiting and classifying participants; Almut Dünnebeil for cognitive assessments; and the participants and their families for their time and interest. This work was supported by the EMPIR programme, co-financed by the Participating States and from the European Union’s Horizon 2020 research and innovation programme, under grant numbers 15HLT04NeuroMET, 18HLT09NeuroMET2, and 22HLT07NEuroBioSTand. Agnes Flöel was supported by the German Research Foundation (327654276-SFB 1315, and 467143400-RU 5429). This study was supported by Topsector Life Sciences & Health (PPP-allowance;#LSHM20106), ABOARD ZonMW, Alzheimer Nederland, Selfridges Group Foundation, Alzheimer’s Drug Discovery Foundation, the Dutch Research Council (#73305095007), Health Holland, Alzheimer’s Association, National Multiple Sclerosis Society, JPND (bPRIDE), EPND (IMI 2 Joint Undertaking) (101034344), Innovative Medicines Initiative 3TR Horizon 2020, grant no 831434, Marie Curie international training network (860197(MIRIADE)). We used large language models, e.g., ChatGPT, during the drafting of the statistical code and to refine the language and clarity of this manuscript. All edits were reviewed and confirmed by the authors.

## CRediT Author Contributions

**Conceptualization** - Laura Göschel; Conceived and designed the study, developed the research question and hypotheses. **Methodology** - Laura Göschel, Ariane Fillmer, Andrea Dell’Orco, Péter Körtvélyessy, Nicolai Franzmeier, Sebastian Römer-Cassiano; Developed and refined the analytical and statistical approach; designed MRI/MRS protocols; discussed conceptual framework and interpretation. **Software** - Andrea Dell’Orco, Ariane Fillmer, Semiha Aydin; Processed fMRI and MRS data; implemented data management and quality control pipelines. **Validation** - Ariane Fillmer, Péter Körtvélyessy; Verified the integrity and quality of MRS data and statistical analyses. **Formal Analysis** - Laura Göschel; Conducted statistical modeling and data analysis. **Investigation** - Laura Göschel, Andrea Dell’Orco, Semiha Aydin, Ariane Fillmer; Acquired and curated MRI/MRS data; coordinated data collection and participant recruitment. **Resources**-Charlotte E. Teunissen, Patty Hoede, Jeanette Melin, Leslie Pendrill, Agnes Flöel, Péter Körtvélyessy; Provided biomarker measurements (p-Tau analyses), NeuroMET memory metric data, funding, and infrastructural support. **Data Curation** - Andrea Dell’Orco, Laura Göschel; Organized, cleaned, and managed the multimodal dataset. **Writing – Original Draft** - Laura Göschel; Wrote the manuscript. **Writing – Review & Editing** - All authors; Reviewed and approved the final manuscript. Visualization - Laura Göschel; Created all figures and tables. **Supervision** - Péter Körtvélyessy, Ariane Fillmer; Provided scientific supervision, guidance, and feedback throughout the project. **Funding Acquisition** - Agnes Flöel, Péter Körtvélyessy; Secured financial and institutional support.

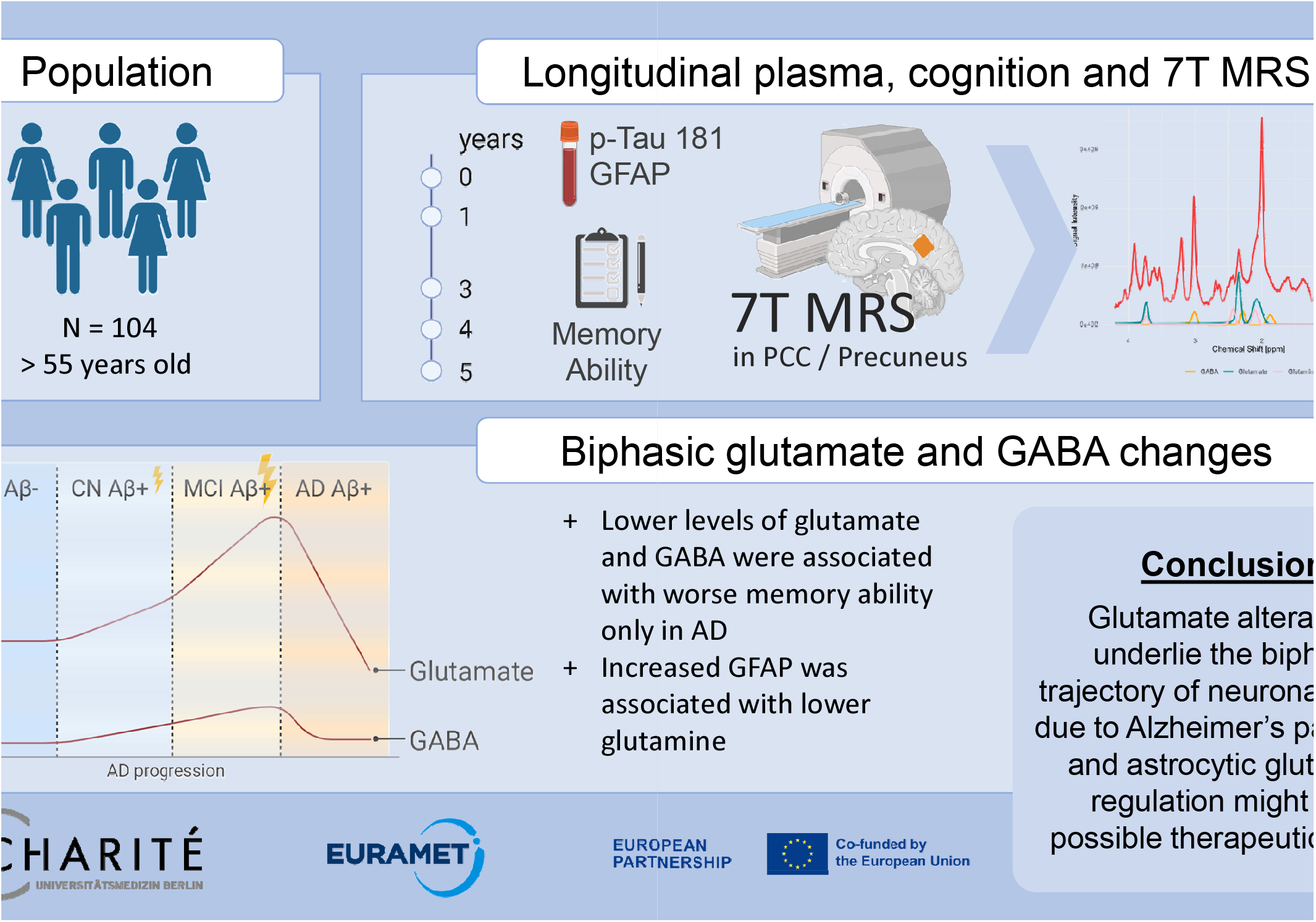

**Extended Figure 1: Longitudinal plasma p-Tau181 increased in the CN Aβ- and AD Aβ+ groups, memory ability decreased in the AD Aβ+ group, and plasma GFAP increased in the CN Aβ-, CN Aβ+ and AD Aβ+ groups.** Displayed β coefficients and p-values indicate estimated annual change per group, based on estimated marginal trends of model 1 (df = 1 for year and age for p-Tau181, memory ability, and plasma GFAP). Amyloid β positivity (Aβ +) was defined as plasma p-Tau181 > 2.08 pg/mL, as indicated by the orange points and dashed line. Abbreviations: AD = Alzheimer’s disease, Aβ = amyloid-beta, CN = cognitively normal, GFAP = glial fibrillary acidic protein, MCI = mild cognitive impairment, NMM = NeuroMET Memory Metric, p-Tau181 = tau phosphorylated at threonine 181.

**Extended Figure 2: Age-related trajectories and sex differences in 7T MRS glutamate, GABA, and their ratio at visit 1.** (A– C) Age at visit 1 had a non-linear effect on the glutamate concentrations with a minimum around 70 years (F(2, 80.57) = 3.21, p = 0.046, model 1), while GABA and the glutamate/GABA ratio exhibited no substantial association. (D–F) Female participants showed lower glutamate levels than males with an estimated difference of -0.78 mmol/L (95% CI = [-1.24; - 0.33], p < 0.001). No sex differences were observed for GABA or the glutamate/GABA ratio. Abbreviations: AD = Alzheimer’s Disease, Aβ+ = Amyloid-beta positive by plasma, CN = Cognitively Normal, GABA = Gamma-Aminobutyric Acid, MCI = Mild Cognitive Impairment, MRS = Magnetic Resonance Spectroscopy.

