## Supplements for "Longitudinal 7T MRS Study of Glutamate and GABA Dynamics in Alzheimer’s Disease Progression: From hyper- to hypoexcitation"

*Table A 1: Degrees of freedom for natural spline terms in all mixed-effects models*

| Predictor | Glutamate | GABA | log(Glutamate/GABA) | Glutamine | log(Glutamate/Glutamine) |
|---|---|---|---|---|---|
| <u>Model 1</u> | | | | | |
| Time | 2 | 1 | 1 | – | – |
| Age | 2 | 1 | 2 | – | – |
| <u>Model 2</u> | | | | | |
| Plasma p-Tau181 | 1 | 1 | 1 | – | – |
| Time | 1 | 1 | 1 | – | – |
| Age | 2 | 1 | 2 | – | – |
| <u>Model 3</u> | | | | | |
| Neurotransmitter | 1 | 1 | 1 | – | – |
| Time | 2 | 2 | 2 | – | – |
| Age | 1 | 1 | 1 | – | – |
| <u>Model 4</u> | | | | | |
| Plasma GFAP | 1 | 1 | 1 | 1 | 1 |
| PCC + Precuneus | 1 | 1 | 1 | 1 | 1 |
| Time | 1 | 1 | 1 | 1 | 1 |
| Age | 2 | 1 | 2 | 2 | 1 |

Note: "–" indicates that the metabolite or ratio was not included in that model family.

### A1 Sensitivity analyses: longitudinal trajectories of groups are non-linear when including one influential data point of the AD group

For transparency, this supplementary analysis reports the longitudinal trajectories of glutamate, GABA, and the log(glutamate/GABA) ratio when a single, influential longitudinal data point is included in the dataset. This data point (highlighted in red in Figure A1 and A2) was identified as highly influential during model fitting. Still, it was retained for this supplementary analysis as no technical, clinical, or quality reason (e.g., medication, comorbidities, or scan quality issues) could justify its removal from the dataset (Figure A1).

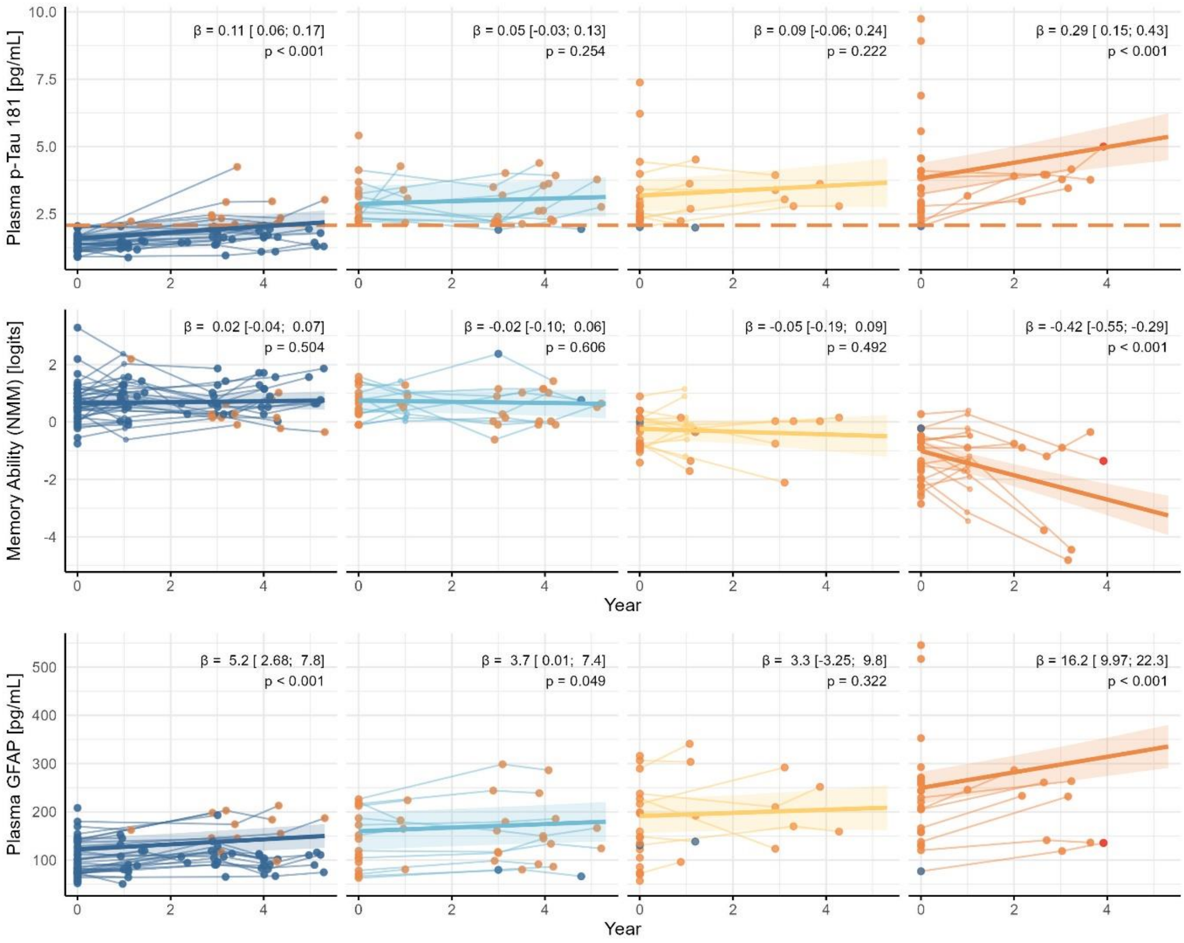


**Figure A1: There was no clinical justification to remove the single influential longitudinal MRS data point in the AD Aβ+ group (red).** Plasma biomarkers and memory ability, measured by NMM, appeared consistent with other data points in the group. Abbreviations: *AD = Alzheimer's disease, Aβ = amyloid-beta, CN = cognitively normal, GFAP = glial fibrillary acidic protein, MCI = mild cognitive impairment, NMM = NeuroMET Memory Metric, p-Tau181 = tau phosphorylated at threonine 181.*

### Glutamate Model Comparison

The influential point significantly impacted the optimal model A1 structure for glutamate:

1. Non-linearity: After comparisons of model fit (ΔAIC = 4.36, ΔBIC = -8.73; $\chi^2(4) = 12.36$, p = 0.015), glutamate trajectory was better fitted to be non-linear across all groups (natural cubic spline df = 2 for years from visit 1), which complicates interpretation and adds complexity to the model where a simpler model may suffice.
2. U-shaped trajectory: The influential point was the only longitudinal observation in the AD group showing an increase over time. Nevertheless, it was sufficient to result in a U-shaped trajectory in the AD group, which does not appear representative of the remaining AD participants.
   - **Main analysis glutamate model** (df=1, influential point removed)
     Glutamate ~ diagnostic group × ns(years from visit 1, df = 1) + ns(age at visit1, df = 2) + sex + (1|record_id)
   - **Sensitivity analysis glutamate model** (df=2, influential point included)
     Glutamate ~ diagnostic group × ns(years from visit 1, df=2) + ns(age at visit1, df=2) + sex + (1|record_id)

### GABA and log(Glutamate/GABA) Models

For the models predicting GABA and the log(Glutamate/GABA) ratio, the presence of the single influential data point did not alter the optimal model structure. Therefore, for all reported supplementary analyses involving GABA and the ratio, the time term was kept at df=1, consistent with the main analyses:

- **GABA model:**
  GABA ~ diagnostic group × ns (years from visit 1, df = 1) + ns (age at visit1, df = 1) + sex + (1|record_id)
- **log(glutamate/GABA):**
  log(glutamate/GABA) ~ diagnostic group × ns(years from visit 1, df=1) + ns(age at visit1, df = 2) + sex + (1|record_id)

### Glutamine and log(Glutamate/Glutamine) Models

Similar to glutamate models, the influential point significantly impacted the optimal model structure for glutamine:

1. Non-linearity: After comparisons of model fit (ΔAIC = 6.38, ΔBIC = -6.75; $\chi^2(4) = 14.34$, $p = 0.006$), glutamine trajectory was better fitted to be non-linear across all groups (natural cubic spline df = 2 for years from visit 1).
2. U-shaped trajectory: The influential point was sufficient to result in a U-shaped trajectory for the AD group that does not seem to be representative of the remaining AD participants.
   - **Main analysis glutamine model** (df=1, influential point removed)
     Glutamine ~ diagnostic group × ns(years from visit 1, df = 1) + ns(age at visit1, df = 2) + sex + (1|record_id)
   - **Sensitivity analysis glutamine model** (df=2, influential point included)
     Glutamine ~ diagnostic group × ns(years from visit 1, df=2) + ns(age at visit1, df=2) + sex + (1|record_id)
1. For the models predicting the log(glutamate/glutamine) ratio, the presence of the single influential data point did not alter the optimal model structure. Therefore, for supplementary analyses involving log(glutamate/glutamine) ratio, the time term was kept at df=1, consistent with the main analyses:
   log(glutamate/glutamine) ~ diagnostic group × ns(years from visit 1, df=1) + ns(age at visit1, df = 1) + sex + (1|record_id)

In summary, the findings suggest that due to missing longitudinal data in the AD group, the group-level patterns are sensitive to the influence of a small number of individuals with extreme trajectories.

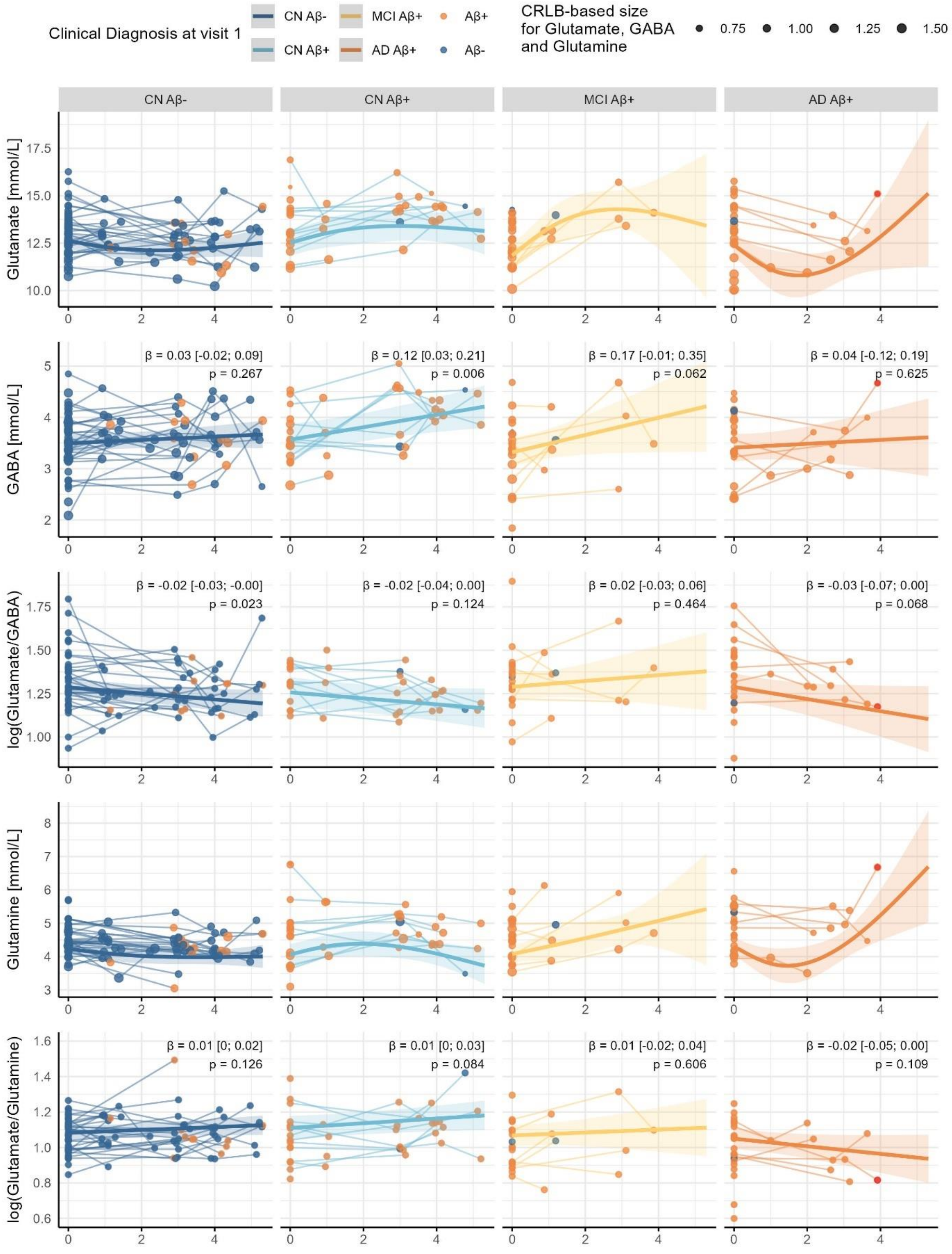


***Figure A 2: Sensitivity analyses of group comparisons of longitudinal changes of 7T MRS neurotransmitter concentrations.*** *The red data point represents an influential point that was considered only in this sensitivity analysis. Longitudinal glutamate and glutamine levels followed a non-linear trajectory with initial increases in the CN Aβ+ and MCI Aβ+ groups, and decreased in the AD Aβ+ group. The one influential data point led to a later increase in the AD Aβ+ group. GABA levels increased substantially over time in the CN Aβ+ and MCI Aβ+ groups. In contrast, the glutamate/GABA ratio declined significantly in the CN Aβ- group and, like the glutamate/glutamine ratio, remained stable for the rest of the groups. All data points were sized using Cramér–Rao lower bounds (CRLB), a common metric for quantifying uncertainty in MRS. Here, smaller datapoints represent higher uncertainty (i.e., higher CLRBs). Amyloid-β positivity (Aβ+) was defined as plasma p-Tau181 > 2.08 pg/mL, as*

*indicated by the orange points. Displayed β coefficients and p-values in GABA, Glutamate/GABA, and Glutamate/Glutamine panels indicate estimated annual change per group, based on estimated marginal trends. Abbreviations: AD = Alzheimer's disease, Aβ = amyloid-beta, CN = cognitively normal, CRLB = Cramér-Rao lower bound, GABA = gamma-aminobutyric acid, MCI = mild cognitive impairment, MRS = magnetic resonance spectroscopy.*

### A2 Longitudinal changes of plasma p-Tau181 were not associated with longitudinal neurotransmitter changes

This supplementary analysis investigated whether individual longitudinal changes in plasma p-Tau 181 influenced the trajectories of MRS concentrations of glutamate, GABA, and log(Glutamate/GABA). For this analysis, we used the dataset excluding the one influential data point in the AD Aβ+ group, which is analogous to the main analysis in the main text.

First, the individual rate of p-Tau181 change (slopes) over time was calculated for each participant using a simple linear model: p-Tau181 slopes = lm(p-Tau181 ~ year).

Next, these calculated pTau181 slopes were incorporated into linear mixed-effects models as a continuous predictor to test interactions with diagnostic group and time. The models allowed for non-linear associations by using natural splines (ns). The df values were chosen based on the best AIC and BIC model fits (df = 2 for the age term in the glutamate and ratio models; the rest, df = 1). Glutamate and GABA models were weighted by Cramér–Rao lower bounds (CRLB), a measure of uncertainty.

**Model A1**:

Neurotransmitter ~ ns(plasma biomarker slopes) × diagnostic group × ns(years from visit 1) + ns(age at visit1) + sex + (1|record_id)

First, these supplementary models indicated that group-wise trajectories of neurotransmitters were not significantly modulated by individual rates of change in plasma p-Tau181. The three-way interaction terms were non-significant for all metabolites:

- Glutamate: $F(3, 97.67) = 0.89$, $p = 0.449$
- GABA: $F(3, 98.78) = 0.74$, $p = 0.529$
- log(Glutamate/GABA): $F(3, 102.4) = 2.11$, $p = 0.104$

Estimated trajectories for individuals with increasing plasma p-Tau181 are shown in Figure A3. Although the interaction terms did not reach statistical significance, the visualization of the glutamate and Glu/GABA ratio models suggests a potential pattern: MCI Aβ+ participants with lower p-Tau181 slopes (presumably reflecting earlier MCI) showed increasing glutamate levels than those with steeper p-Tau181 increases (presumably more advanced MCI). This trend is consistent with the hypothesis that rising plasma p-Tau181 is associated with glutamatergic dynamics in early AD pathology. However, the limited longitudinal sample size in the MCI group likely reduced statistical power, preventing these effects from reaching significance.

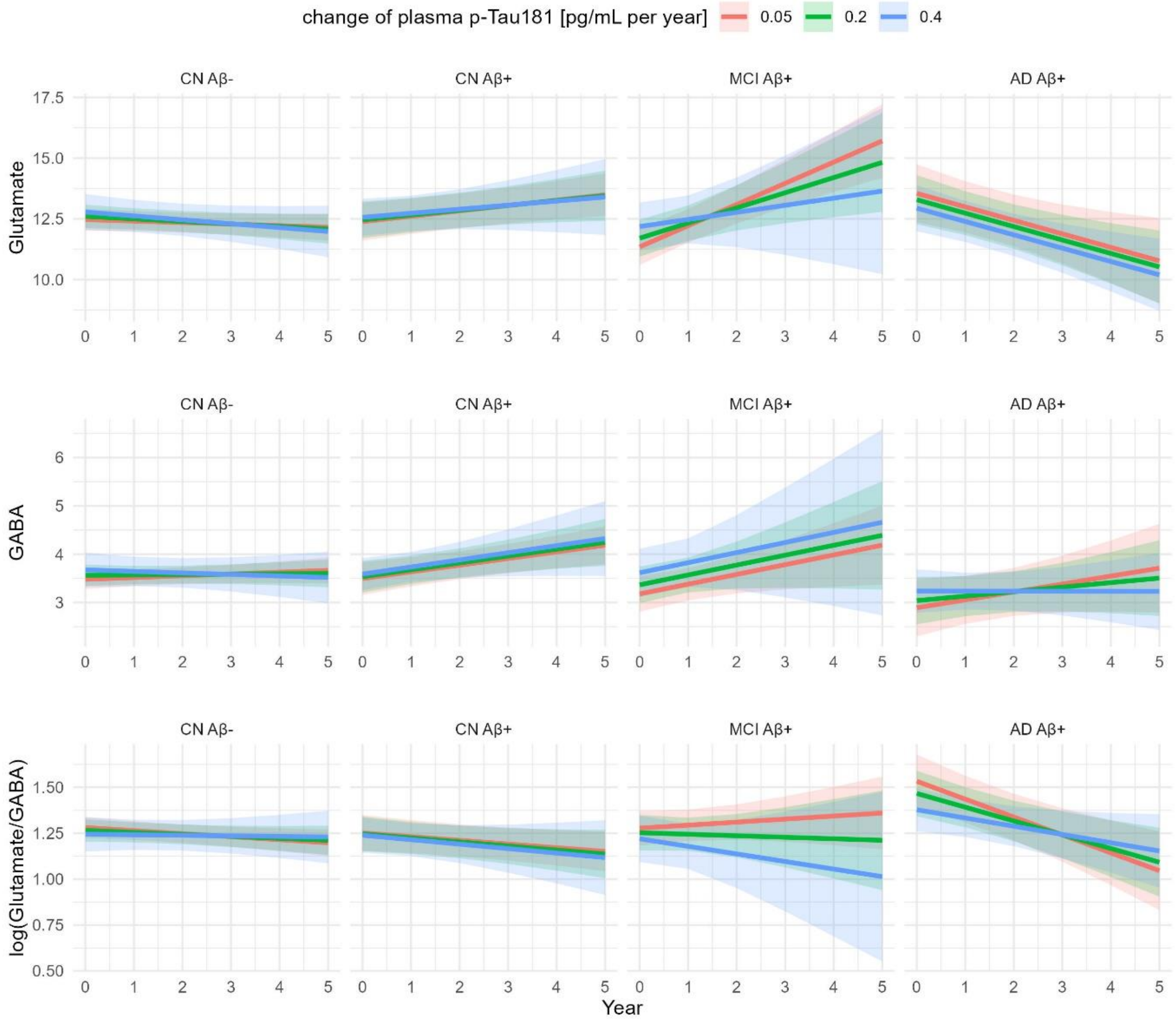


***Figure A 3: Estimated group-wise longitudinal trajectories of MRS glutamate, GABA, and log(glutamate/GABA) for individuals with increasing plasma p-Tau181.*** *The data are shown by diagnostic group. Within each panel, the trajectories are visualized for three representative p-Tau181 slopes as indicated in the legend. The lines represent the estimated marginal means from the linear mixed-effects models, and the shaded areas represent the 95% confidence intervals. Statistical analysis confirmed that the neurotransmitter trajectories were not significantly influenced by the individual p-Tau181 slopes, as indicated by the non-significant three-way interaction term (pTau slopes × diagnostic group × year) across all three metabolites (glutamate: $p = 0.449$; GABA: $p = 0.529$; log(glutamate/GABA): $p = 0.104$). Abbreviations: AD = Alzheimer's disease, Aβ = amyloid-beta, CN = cognitively normal, GABA = gamma-aminobutyric acid, MCI = mild cognitive impairment, MRS = magnetic resonance spectroscopy, p-Tau 181 = tau phosphorylated at threonine 181.*

### A3 Longitudinal changes of plasma GFAP were not associated with longitudinal neurotransmitter changes

This supplementary analysis investigated whether individual longitudinal changes in plasma GFAP influenced the trajectories of MRS concentrations of glutamate, GABA, glutamine, and log(glutamate/glutamine). For this analysis, we used the dataset excluding the one influential data point in the AD Aβ+ group, which is analogous to the main analysis in the main text.

First, the individual rate of GFAP change (slopes) over time was calculated for each participant using a simple linear model: GFAP slopes = lm(GFAP ~ year).

Next, we used model A1 described in section A2, and df values were chosen based on the best AIC and BIC fits (df = 2 for the years term in the glutamine model and for the age term in the glutamate model; df = 1 for the rest).

The supplementary models and their visualization in Figure A4 indicated that individual rates of change in plasma GFAP did not significantly modulate group-level trajectories of neurotransmitters. The three-way interaction terms were non-significant for all metabolites:

- Glutamate: $F(3, 105.93) = 0.67$, $p = 0.572$
- GABA: $F(3, 114.51) = 0.37$, $p = 0.776$
- Glutamine: $F(6, 87.01) = 1.69$, $p = 0.134$
- Log(Glutamate/Glutamine): $F(3, 101.93) = 1.28$, $p = 0.286$

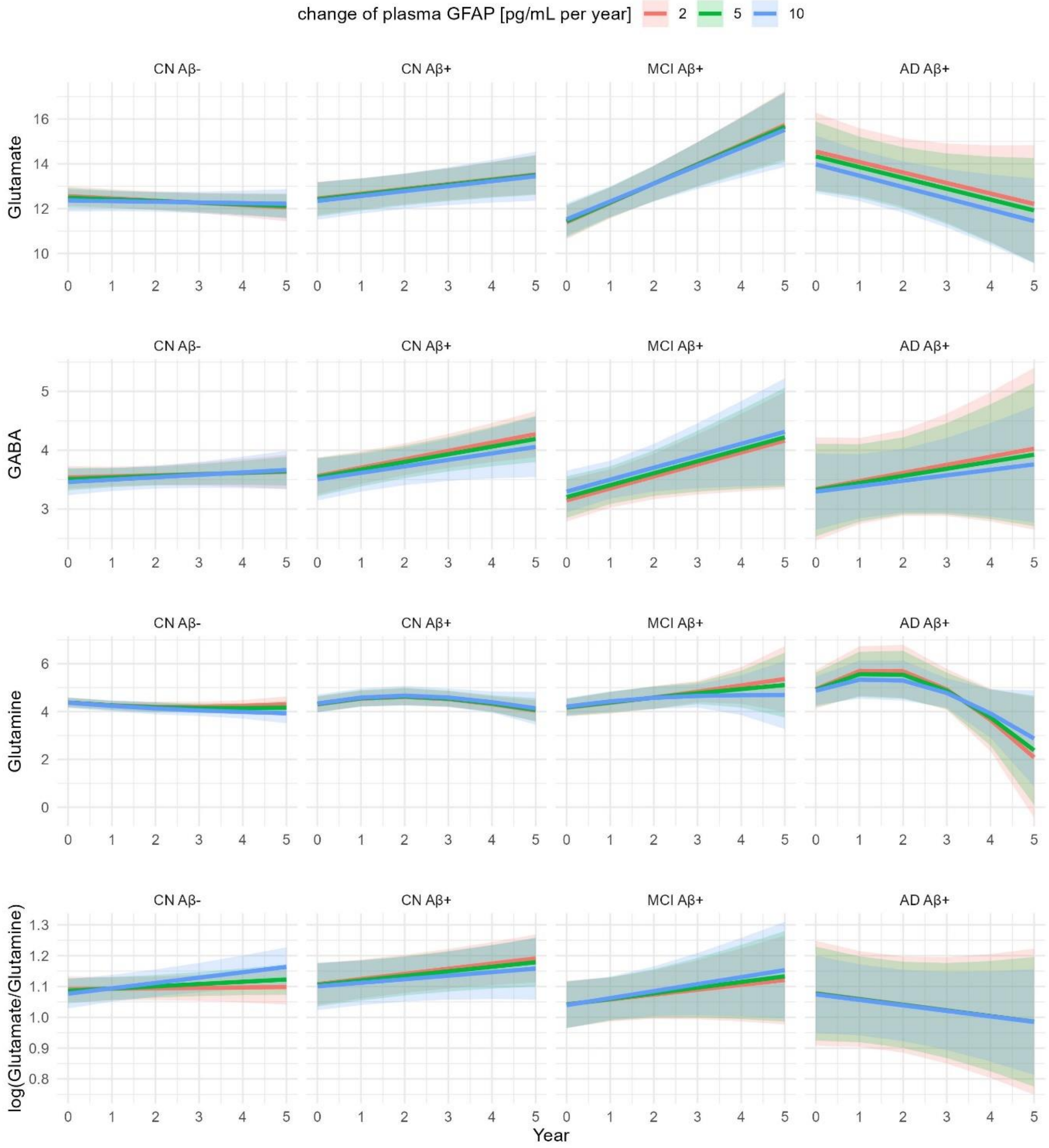

*Figure A 4:* ***Estimated group-wise longitudinal trajectories of MRS glutamate, GABA, glutamine, and log(glutamate/glutamine) for individuals with increasing plasma GFAP.*** *The data are shown by diagnostic group. Within each panel, trajectories are visualized for three representative GFAP slopes, as indicated in the legend. The lines represent the estimated marginal means from the linear mixed-effects models, and the shaded areas represent the 95% confidence intervals. Statistical analysis confirmed that the neurotransmitter trajectories were not significantly influenced by the individual GFAP slopes, as indicated by the non-significant three-way interaction term (GFAP slopes × diagnostic group × year) across all metabolites (glutamate: p = 0.572; GABA: p = 0.776; glutamine: p = 0.134; log(glutamate/glutamine): p = 0.286). Abbreviations: AD = Alzheimer's disease, A$\beta$ = amyloid-beta, CN = cognitively normal, GABA = gamma-aminobutyric acid, GFAP = glial fibrillary acidic protein, MCI = mild cognitive impairment, MRS = magnetic resonance spectroscopy.*
