## Supplementary figures and images for "Longitudinal 7T MRS Study of Glutamate and GABA Dynamics in Alzheimer’s Disease Progression: From hyper- to hypoexcitation"

### extended figure 1

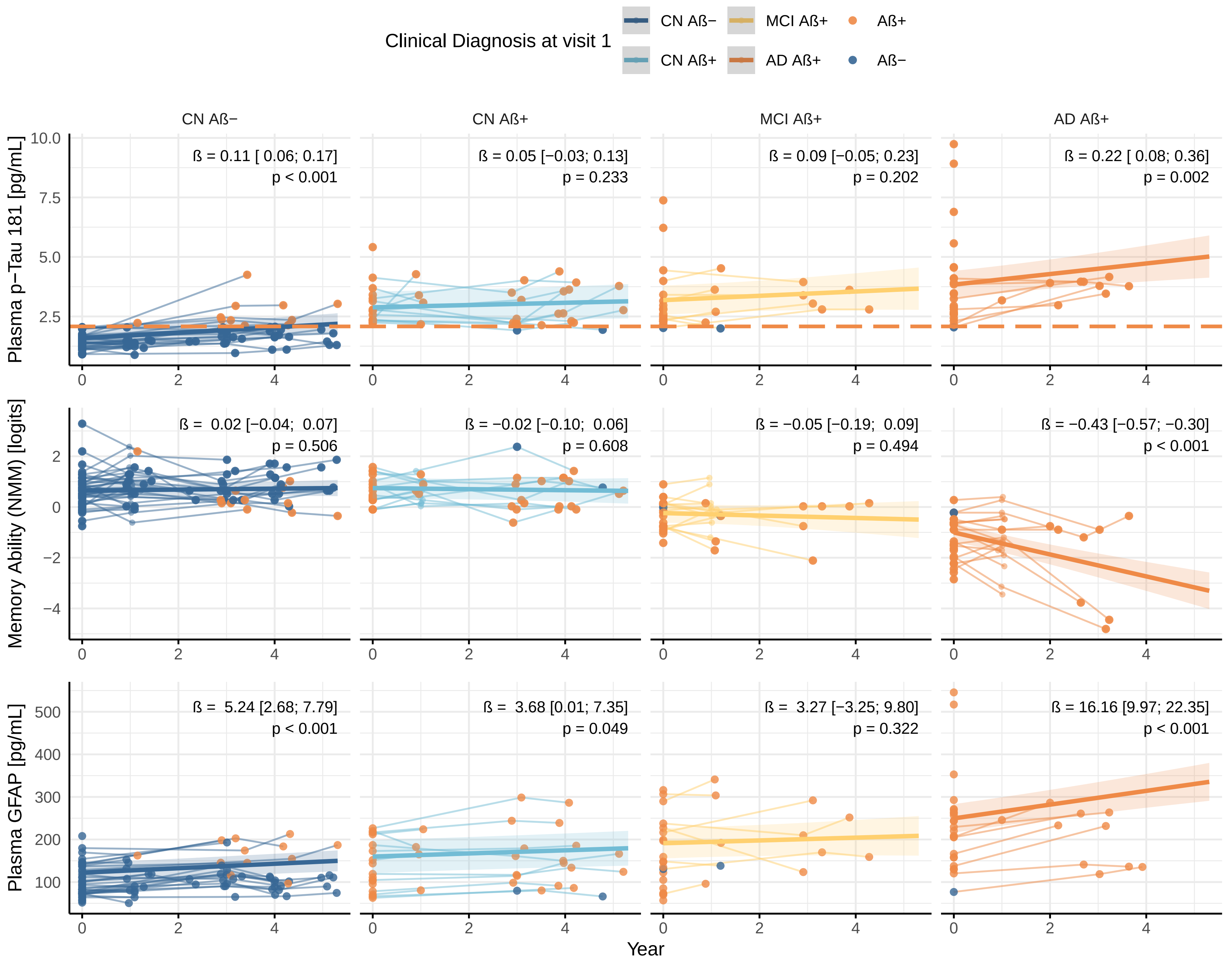

### extended figure 2

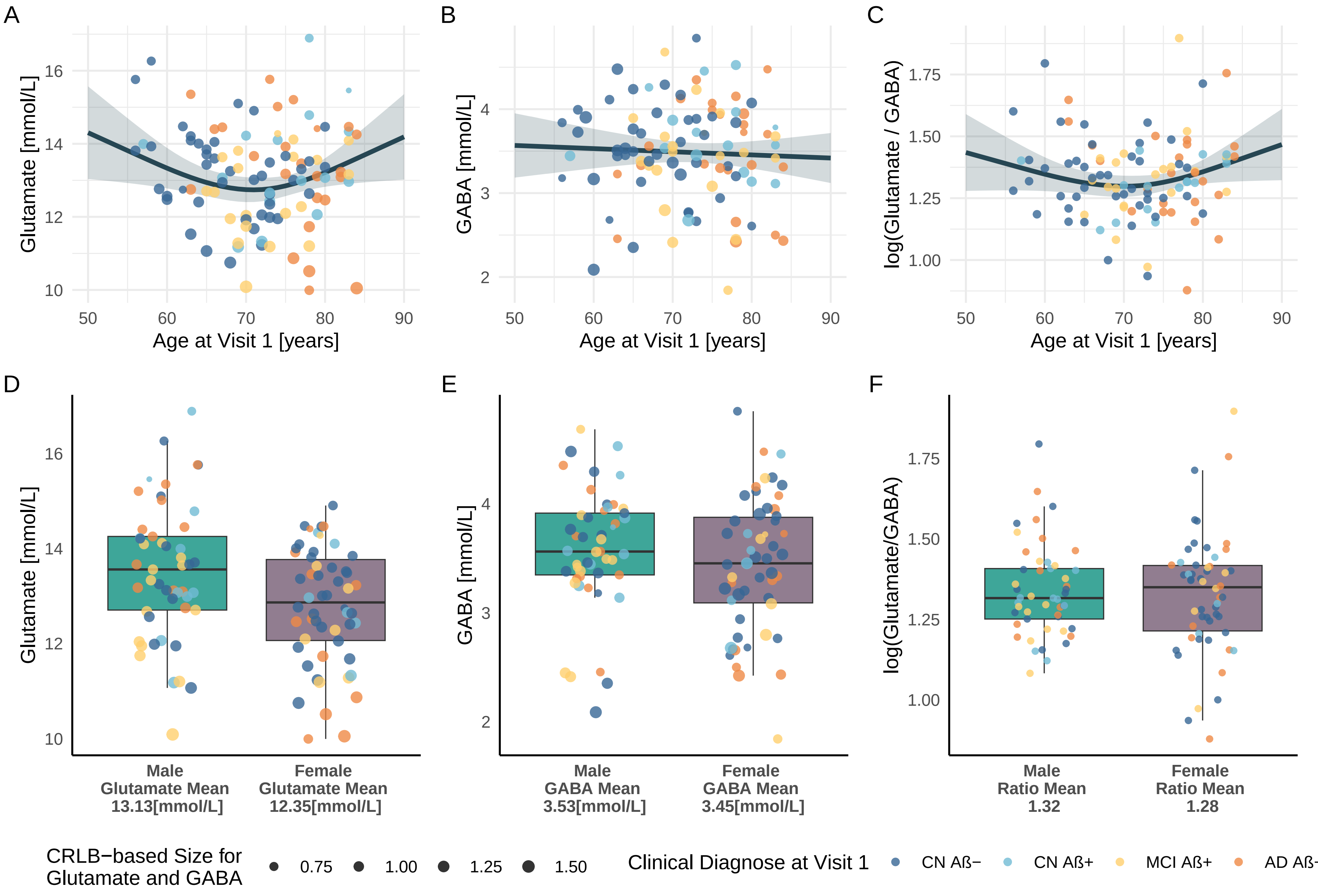
